# Delayed Retention of Technical Surgical Skills Following Virtual, Augmented, and Mixed-Reality Simulation: A Systematic Review and Meta-analysis

**DOI:** 10.64898/2026.09.15.26363133

**Authors:** Faraz Shamim, Raziya Akhtar Hussain

## Abstract

**Background:** Extended-reality simulation is widely used for technical surgical training, yet immediate gains do not establish that skills persist once practice stops.

**Objective:** To characterize delayed technical-skill outcomes after virtual, augmented, or mixed-reality training and estimate the comparative effect at the first eligible uncontaminated delayed assessment.

**Methods:** PubMed/MEDLINE, Europe PMC, CENTRAL, ERIC, and ClinicalTrials.gov were searched from inception through May 5, 2026. Eligible studies enrolled surgical learners or practitioners, used virtual, augmented, or mixed reality for technical training, and measured objective performance at least seven days later. Pure retention was distinguished from maintenance and reacquisition. One endpoint per independent study family was selected. Standardized mean differences were expressed as Hedges’ g, oriented so positive values favored extended reality. Paule-Mandel random effects with Hartung-Knapp confidence intervals were primary.

**Results:** Thirty-four reports representing 33 studies (1,186 participants; 21 randomized trials) from 12 countries were included. Six studies (162 participants in selected arms) were pooled. The summary effect was g = 1.39 (95% CI -0.94 to 3.72; I² = 88.0%; τ² = 4.53); the 95% prediction interval was -5.04 to 7.81. Excluding the figure-derived estimate reduced the effect to g = 0.61 (95% CI -0.76 to 1.99). GRADE certainty for the primary outcome was very low. No pooled study assessed an interval of 90 days or longer.

**Conclusions:** Delayed technical advantages after extended-reality simulation were observed in some settings, but the comparative evidence does not define a precise or generalizable retention effect. Substantial heterogeneity, sparse transfer and longer-term evidence, comparator differences, and influence from one figure-derived estimate warrant cautious interpretation.

## Introduction

Simulation addresses the tension between safe patient care and the need for surgeons to acquire complex motor skills. Reduced operative exposure, variable case mix, duty-hour constraints, and closer scrutiny of outcomes leave less room for unstructured repetition. Simulators permit deliberate practice and errors without patient harm while standardizing task difficulty and feedback. Meta-analytic and randomized evidence supports simulation for technical performance, including transfer to the operating room in selected settings [1–3]. Recent reviews likewise find that digital and extended-reality tools can support surgical skill acquisition, while emphasizing heterogeneous methods and limited evidence on durable, clinically proximal outcomes [6,7]. However, evidence of acquisition does not establish how long an advantage remains after structured practice ends.

Extended-reality technologies broaden this environment. Virtual reality can present repeatable procedural scenarios and quantify time, motion, errors, and task completion; augmented reality can place digital guidance within a physical task; and mixed reality can couple spatially anchored content with the surrounding environment. Learners may receive visual, haptic, or automated performance feedback and practice outside clinical scheduling. Platforms vary in immersion, physical fidelity, feedback, and correspondence with the eventual clinical task, so their educational effect depends on the practice conditions they create rather than the XR label alone [8,9].

Most evaluations emphasize acquisition curves or immediate post-training performance. These endpoints may reflect recent feedback, repeated exposure to identical cues, and temporary familiarity with a device’s controls or scoring logic. A learner can therefore show a large immediate gain without retaining the same advantage after disuse. For educators deciding whether training supports readiness weeks or months later, durability is distinct from the post-test peak.

Delayed performance reflects the interaction of consolidation and decay. Practice can stabilize a motor representation after training, but later accessibility depends on initial learning, task complexity and specificity, prior experience, subsequent exposure, and interference. Procedural skills may decline at different rates, and loss from a post-training peak can occur even when performance remains above baseline [4,5]. Apparent stability may instead reflect unmeasured practice during the interval. A delayed endpoint therefore needs a defined interval and an account of intervening exposure. The present review used a minimum delay of seven days while recognizing that a one-week and a six-month assessment address different educational horizons; evidence on spacing also supports treating practice schedule as a material design feature [10].

Same-task retention and transfer also represent different questions. Repeating the trained simulator task tests whether a learner can reproduce a performance pattern under familiar perceptual cues, device mechanics, and scoring rules. Transfer testing changes one or more of those conditions by using another simulator, a physical model, cadaveric or animal tissue, or an operative task. It asks whether the learner can adapt the underlying technical skill when the environment no longer reproduces the training interface. Same-task persistence is educationally relevant for documenting memory of the trained task, but transfer is more directly related to whether simulation preparation generalizes beyond the platform. A training program may produce strong same-task retention yet little advantage on a clinically proximal transfer assessment; the two outcomes should therefore not be combined conceptually or interpreted as interchangeable evidence.

Pure retention is delayed performance measured before any relevant booster, refresher, or retraining exposure. Maintenance studies test whether planned intervening practice preserves a standard, whereas reacquisition studies measure how efficiently performance returns after practice resumes. Faster reacquisition may be valuable even when unassisted performance has declined, but it does not show that performance remained intact. Separating these constructs clarifies whether a program produces durable performance, prevents decay through scheduled practice, or facilitates later relearning.

XR-specific synthesis is needed because standardized scenarios, granular process metrics, and device-generated feedback may strengthen both learning and interface familiarity. Recent reviews have evaluated broad XR effectiveness, digital training tools, robotic-simulation transfer, head-mounted XR, and practice spacing [6–10], but none resolves the delayed comparative effect defined here across retention constructs. We therefore reviewed objective technical performance measured at least seven days after XR surgical training, distinguished pure retention from maintenance and reacquisition, separated same-task persistence from transfer, and synthesized independent comparative effects when data permitted. We also examined task relation, comparator type, follow-up interval, statistical method, figure-derived data, and risk of bias.

## Methods

### Study design and reporting

This systematic review and meta-analysis was conducted and reported in accordance with PRISMA 2020 [11]. The review evaluated objective surgical technical performance measured at least seven days after completion of the initial XR training episode.

### Protocol and registration

This systematic review was not prospectively registered on PROSPERO; however, the study protocol was fully established and finalized on May 5, 2026 prior to the commencement of data extraction and analysis.

### Eligibility criteria

Studies were eligible if they enrolled surgical learners or practitioners; evaluated virtual, augmented, or mixed reality as a technical training exposure; and reported an objective technical assessment at least seven days after training. Randomized trials, nonrandomized comparative studies, and prospective longitudinal cohorts were eligible. Knowledge, confidence, satisfaction, presence, and other nontechnical outcomes were excluded. A poolable comparator was not required for inclusion in the systematic review.

### Information sources and search strategy

PubMed/MEDLINE, Europe PMC, the Cochrane Central Register of Controlled Trials (CENTRAL), ERIC, and ClinicalTrials.gov were searched from inception through May 5, 2026. Broad searches combined extended-reality, simulation, surgical or procedural training, and technical-performance concepts. Supplementary retention-focused searches were used to identify delayed outcomes not captured by the broad strategies. Complete source-specific strategies, limits, and retrieval counts are provided in Supplementary Table S1.

### Study selection

All 5,055 deduplicated records underwent title and abstract screening. Prespecified criteria concerning record type, population, intervention, surgical and XR relevance, objective technical outcomes, and delayed-retention relevance were applied manually. Two reviewers independently screened titles and abstracts and assessed potentially eligible full-text reports; disagreements were resolved by discussion and consensus. Of 5,055 screened records, 4,122 were excluded and 933 reports were sought and retrieved for full-text assessment. Full-text exclusions were categorized by failure to meet prespecified population, intervention, outcome, design or report-type, or follow-up criteria.

### Data extraction

One reviewer extracted study and outcome data, and a second reviewer independently verified each entry against the primary report. Discrepancies were resolved by consensus.

### Retention and task classification

Pure retention was defined as objective delayed performance measured before any relevant task-specific booster, refresher, or retraining exposure. Maintenance involved planned practice during the retention interval or immediately before the delayed endpoint. Reacquisition measured performance after retraining began. Follow-up intervals were grouped as short (7–29 days), intermediate (30–89 days), or longer term (at least 90 days). Assessments were classified as same task when the trained simulator and task were repeated, transfer when performance was tested in another environment or procedure, and both when a study reported each form.

### Study-family linkage

Reports sharing participants, recruitment dates, intervention groups, and outcome schedules were treated as one study family. Only one statistically independent contrast from a study family entered any meta-analysis unless a design provided independent participant groups.

### Risk-of-bias assessment

Randomized trials were assessed with RoB 2, nonrandomized comparative studies with ROBINS-I, and nine uncontrolled longitudinal studies with JBI design-specific checklists. Five descriptive retention or validation cohorts were assessed with the JBI cohort checklist, while four one-group before-after or repeated-measures intervention studies were assessed with the JBI quasi-experimental checklist. Tool-specific judgments were retained on their original scales. JBI responses were reported item by item and were not converted into an overall low/moderate/high rating. Two reviewers independently assessed risk of bias, and disagreements were resolved by discussion and consensus.

### Effect-size calculation

The primary estimand was the standardized comparative difference in objective delayed technical performance after XR training at the first eligible uncontaminated delayed assessment. The broad synthesis pooled across task relation and comparator intensity when the endpoint represented delayed surgical technical performance; subgroup analyses examined these sources of clinical heterogeneity. The endpoint hierarchy was defined before pooled effect calculation: a validated composite or global technical score, then a prespecified primary objective metric, procedural quality, errors, economy of motion, and time. When outcomes were otherwise equivalent, the highest-complexity eligible module was selected; ties were resolved by the report’s stated primary outcome and then the most complete variance data. The first delayed assessment at least seven days after training and before any relevant booster, warm-up, refresher, or retraining was preferred. No post hoc exception based on significance was allowed. Hedges’ g was oriented so positive values favored XR, with signs reversed for lower-is-better outcomes. Studies were not pooled when dispersion or compatible quantitative summaries could not be established reliably.

### Statistical analysis

The primary synthesis used a random-effects model with Paule-Mandel estimation of between-study variance and Hartung-Knapp confidence intervals because few heterogeneous studies were expected [12]. Heterogeneity was summarized with Q, I², and τ², and a 95% prediction interval was calculated. Analyses were implemented in R version 4.6.1 (R Foundation for Statistical Computing, Vienna, Austria) with metafor version 5.0-1; the primary model used rma.uni with method=“PM” and test=“knha”. Formal small-study-effect tests were not performed because fewer than 10 studies were pooled. Certainty for the primary outcome was assessed with GRADE across risk of bias, inconsistency, indirectness, imprecision, and publication bias [13].

### Sensitivity and subgroup analyses

For multi-arm studies, one contrast was selected when it best isolated the eligible XR comparison. Independent strata were combined with standard aggregate mean and variance formulas before effect calculation, as for Sabench Pereferrer. No shared control entered more than one primary contrast, no control group was split, and no correlation adjustment was required because the primary model contained one manually verified two-arm contrast per study family.

Exploratory subgroups considered same-task versus transfer assessment, active versus inactive comparator, and short versus intermediate follow-up. Sensitivity analyses used DerSimonian-Laird estimation with a normal confidence interval, restricted the synthesis to randomized trials, excluded the figure-derived estimate, excluded studies at high or serious risk of bias, and omitted each study in turn. Subgroup estimates were interpreted descriptively because the cells were small and task relation was correlated with comparator type.

## Results

### Study selection

The searches identified 8,628 records. After removal of 3,573 duplicates, all 5,055 deduplicated records underwent title and abstract screening; 4,122 were excluded and 933 reports were sought for retrieval. All 933 reports were retrieved and assessed in full text, and 899 did not meet the prespecified eligibility criteria. Thirty-four reports representing 33 distinct studies were included, and six independent studies contributed to the primary meta-analysis (Figure 1).

**Figure 1.**
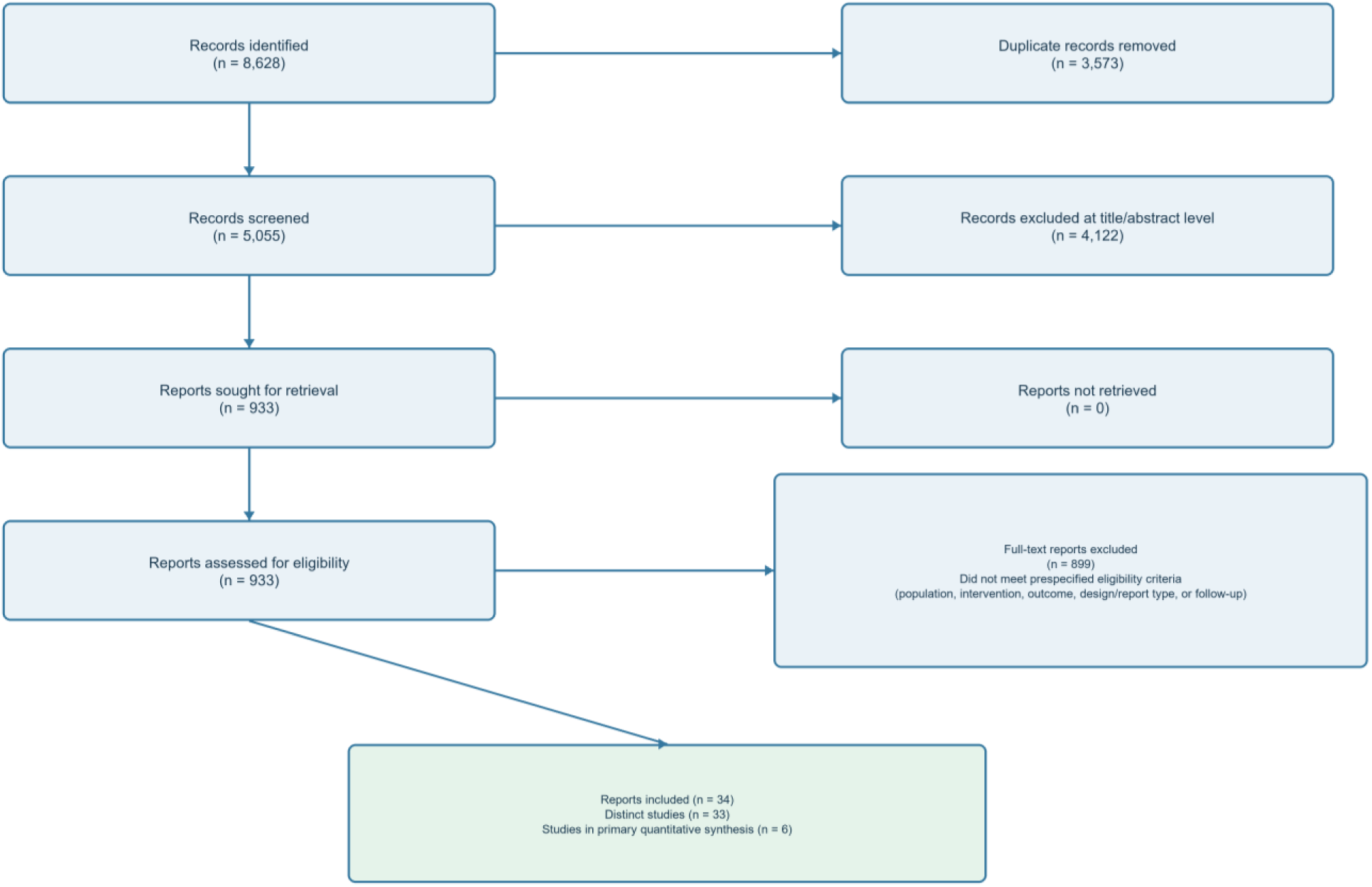
PRISMA 2020 flow diagram. All 5,055 deduplicated records underwent title and abstract screening; 4,122 were excluded. All 933 reports sought for retrieval were obtained and assessed in full text; 899 did not meet the prespecified eligibility criteria.

### Study characteristics

The 33 studies were published from 2004 through 2024 and enrolled 1,186 participants across 12 countries (Table 1) [14–47]. Twenty-one were randomized trials, three were nonrandomized comparative studies, and nine were uncontrolled longitudinal studies. Thirty-one evaluated virtual reality alone, one combined virtual reality with a physical video trainer, and one evaluated augmented reality; no eligible mixed-reality study was identified. The Cychosz 2018 and 2019 reports arose from the same 43-participant cohort and were counted as one study family and one independent meta-analysis contribution [34,35].

**Table 1.**
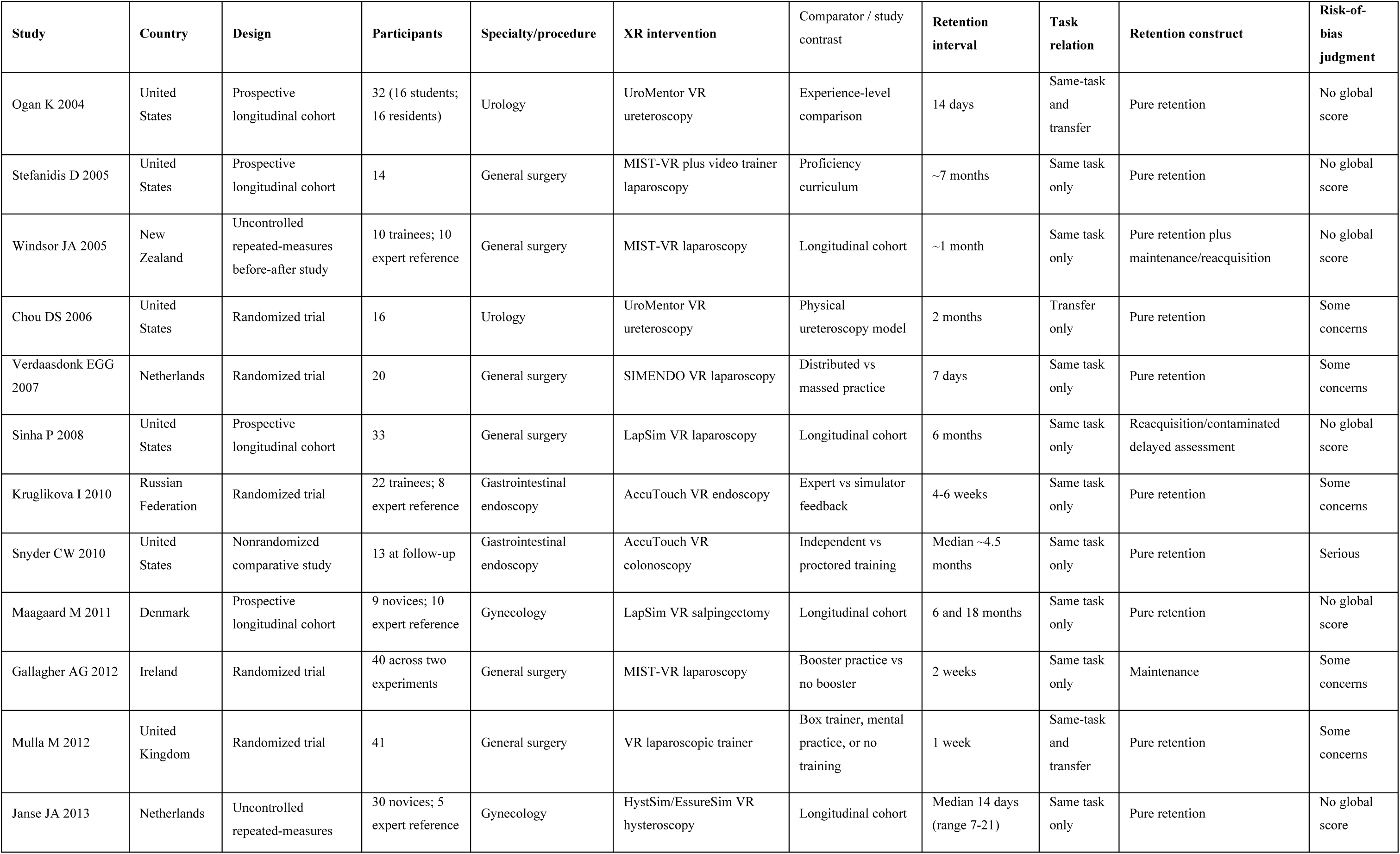
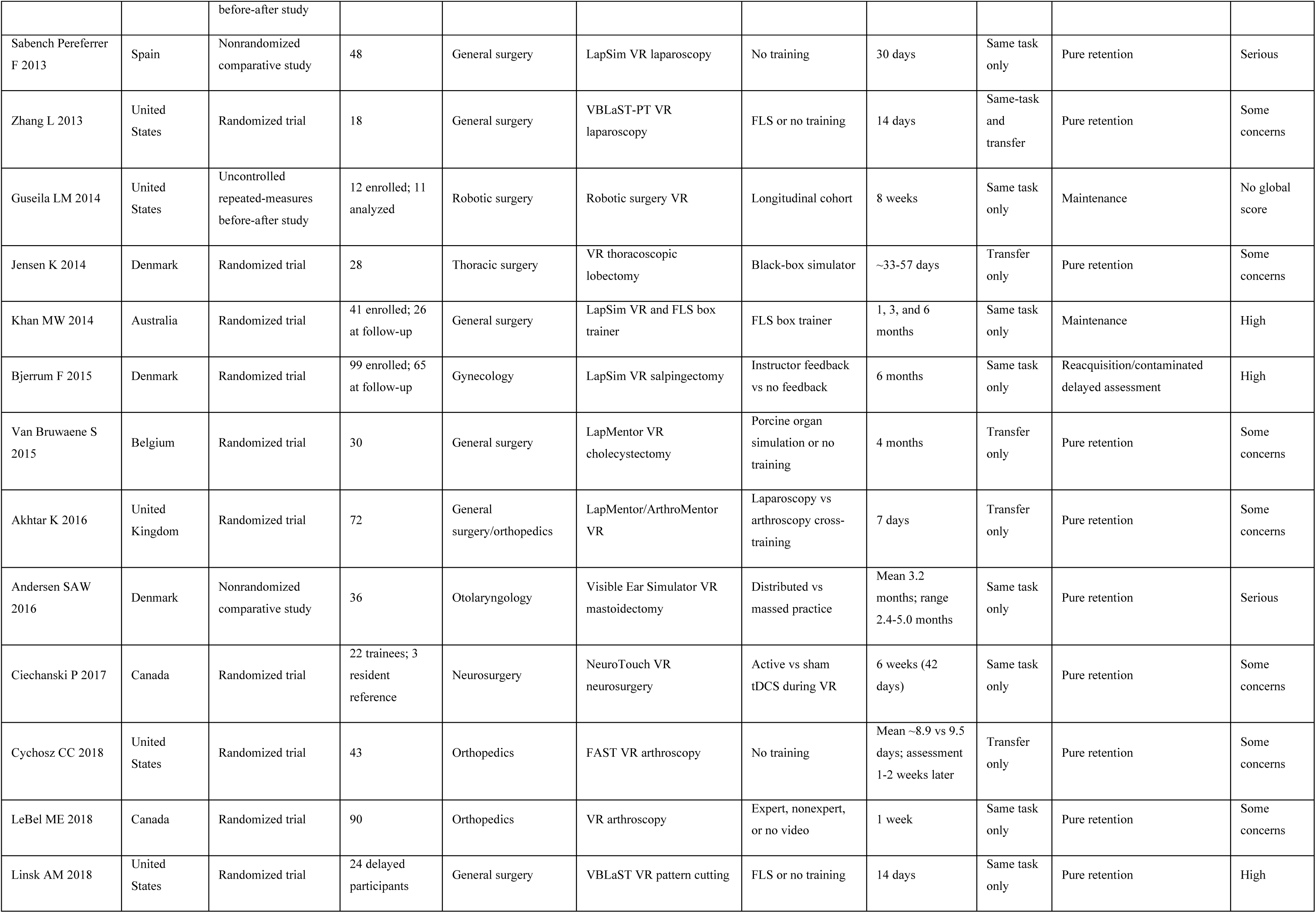
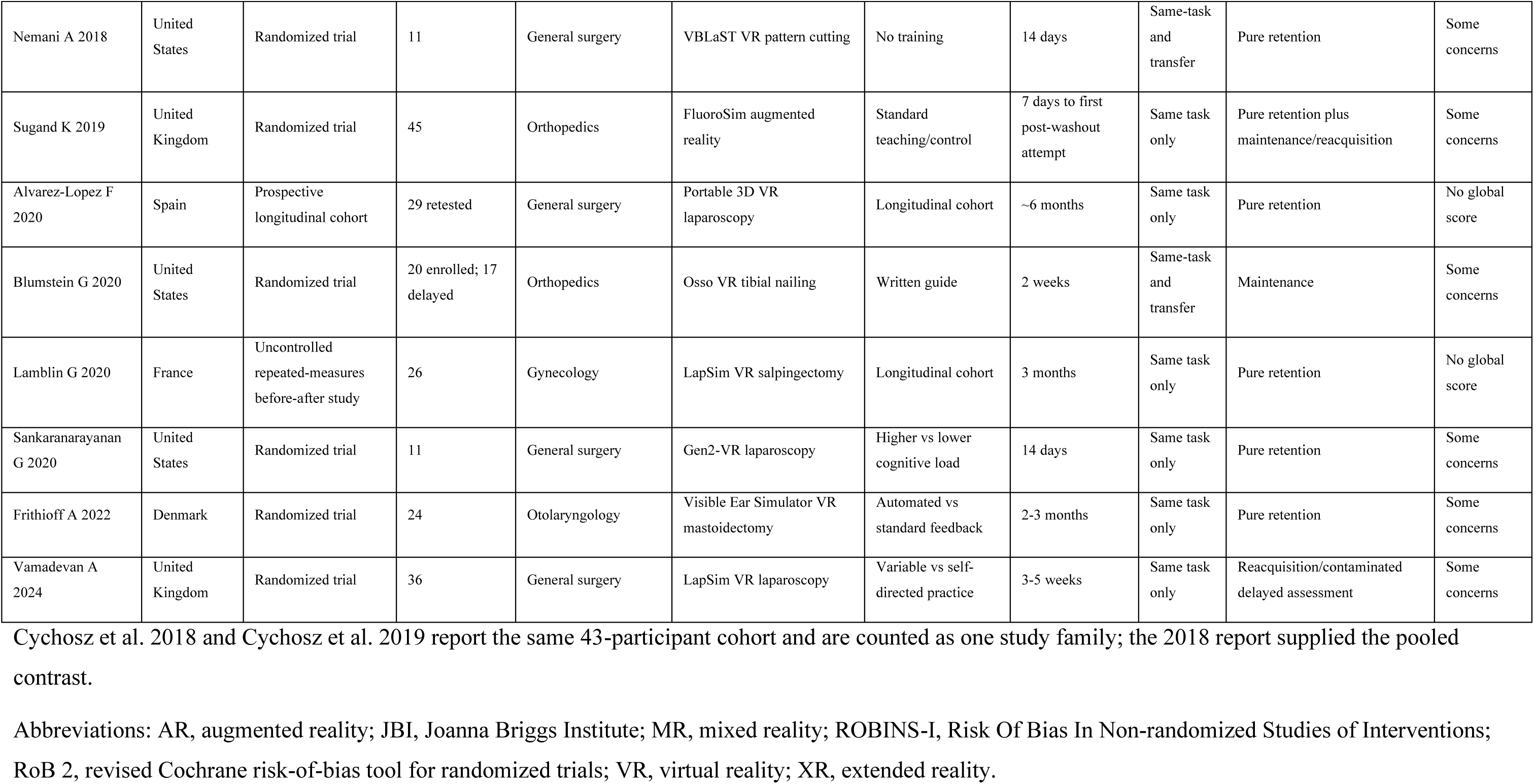
Characteristics of included studies.

Studies addressed basic and procedural laparoscopy, ureteroscopy, gastrointestinal endoscopy, arthroscopy, thoracoscopic lobectomy, mastoidectomy, robotic surgery, neurosurgical skills, and fluoroscopic guidewire placement [14–47]. Participants were predominantly students, residents, registrars, or novices; several studies used experienced surgeons as proficiency references rather than causal comparators. Training ranged from brief supervised sessions to practice until proficiency. Delayed comparisons included no-practice controls, physical trainers or anatomical models, alternative simulator schedules, feedback or supervision strategies, and uncontrolled within-participant follow-up.

The balance of modalities was also uneven. The literature was overwhelmingly based on screen-based or haptic virtual-reality systems, while augmented reality contributed only one eligible study and mixed reality contributed none. Consequently, the review’s quantitative result should be understood primarily as evidence about virtual-reality surgical simulation rather than as equally weighted evidence across all XR modalities. The included technologies also changed substantially over the two decades covered by the review. Older systems often emphasized isolated psychomotor metrics, whereas later studies more often used procedural modules, physical transfer tasks, or combined performance measures. Those differences in interface, feedback, and outcome construction form part of the clinical heterogeneity represented by the pooled estimate.

### Retention patterns

Twenty-four studies evaluated pure retention only, and two included both an uncontaminated retention assessment and maintenance or reacquisition. Four studies evaluated scheduled maintenance, while three assessed reacquisition or delayed performance after additional exposure. Fourteen studies had short follow-up only, six intermediate follow-up only, seven longer-term follow-up only, two spanned short and intermediate follow-up, three spanned intermediate and longer-term follow-up, and one did not permit precise window assignment. Retention intervals ranged from seven days to 18 months, but longer follow-up did not usually coincide with a randomized comparative design. The long-term evidence therefore described whether performance persisted within small cohorts more often than whether XR produced a durable advantage over another educational approach.

Twenty-three studies assessed the same task only, five assessed transfer only, and five reported both. Same-task assessments commonly repeated the trained simulator module or closely related platform metrics. Transfer assessments used another simulator, a physical or anatomical model, cadaveric or animal tissue, or a procedural task outside the original interface. Comparator choice tracked task relation in important ways: inactive or no-training comparators were common in same-task studies, whereas transfer studies more often used an active physical trainer, model, or alternative curriculum. Other studies compared XR strategies with one another, including differences in practice spacing, instructor feedback, observation, cognitive loading, or booster exposure. These designs were informative about how XR might be delivered, but they did not estimate the same contrast as XR training versus no XR training.

### Risk of bias

Among 21 randomized trials, 18 were judged to have some concerns and three were judged at high risk; none was judged low risk overall. RoB 2 judgments were determined with the tool algorithm. High-risk judgments reflected substantial or demonstrably informative missing delayed outcome data. Recurring concerns included incompletely described allocation procedures, attrition between training and delayed testing, and uncertainty about selection among several available outcomes or time points.

All three nonrandomized comparative studies were judged at serious risk, chiefly because of confounding or selection into delayed follow-up. The JBI cohort checklist was used for five descriptive retention or validation cohorts, and the JBI quasi-experimental checklist was used for four one-group intervention studies. Appraisals remained item-level without a global rating (Figure 3). Principal limitations included absence of a concurrent causal comparator, incomplete control of intervening exposure, and uncertain follow-up completeness. Complete item-level judgments are provided in the supplement.

### Primary meta-analysis

Of 33 studies, six provided an independent comparative pure-retention endpoint with sufficient and compatible quantitative data for the primary standardized-effect synthesis [17,26,27,33,34,37]. The remaining studies contributed to the narrative evidence map but were not considered lower quality merely because their designs, constructs, comparators, timing, or reported statistics did not support this estimand. Individual pooled effects ranged from g = -0.78 to 5.86 (Table 2; Figure 2). The Paule-Mandel/Hartung-Knapp summary was g = 1.39 (95% CI -0.94 to 3.72). Heterogeneity was considerable (Q = 41.58, 5 df, p < 0.001; I² = 88.0%; τ² = 4.53), and the 95% prediction interval was -5.04 to 7.81.

**Figure 2.**
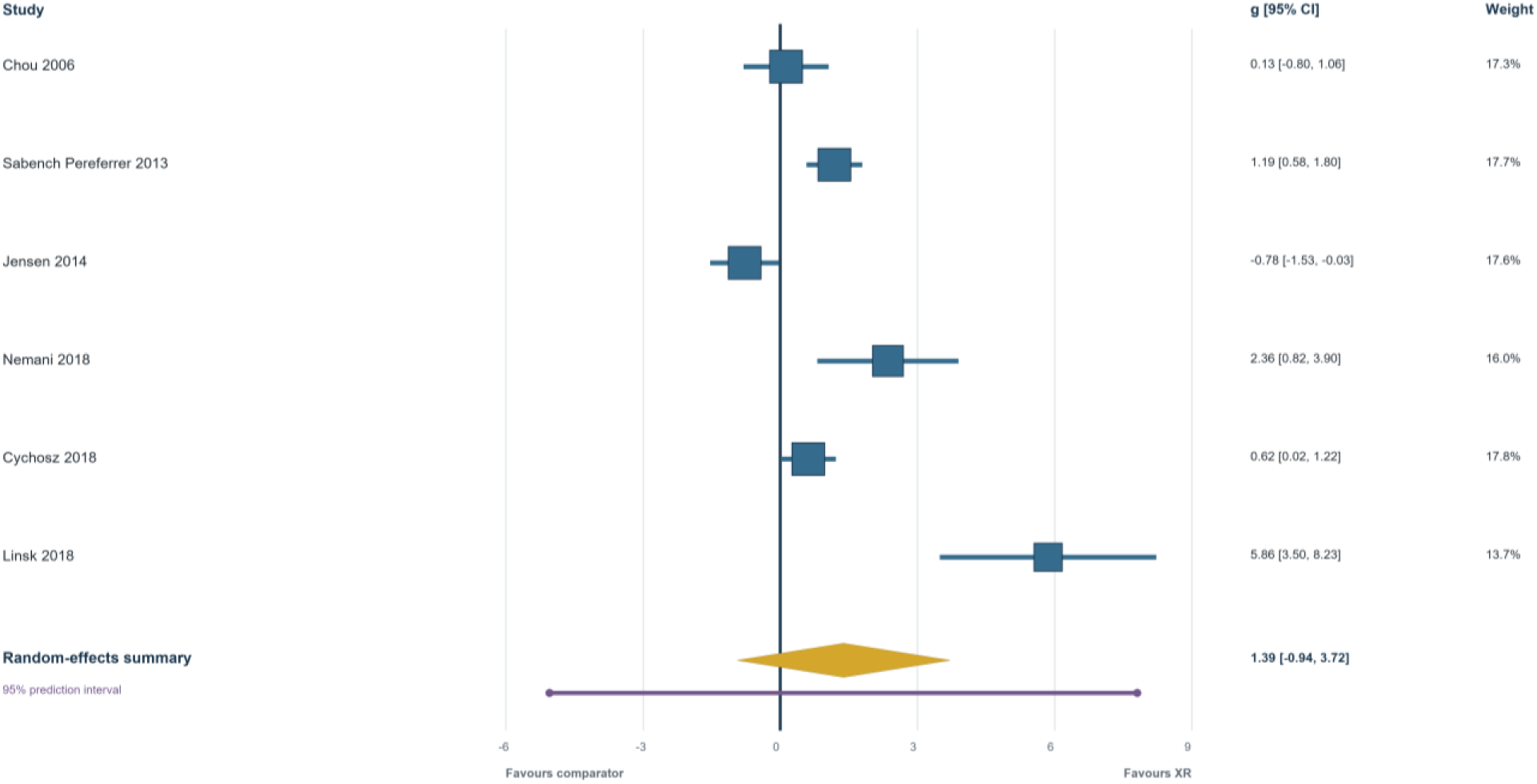
Primary random-effects meta-analysis of delayed objective technical performance. Squares are proportional to Paule-Mandel random-effects weights; horizontal lines show study-level 95% confidence intervals. The diamond shows the Hartung-Knapp summary confidence interval, and the purple line shows the 95% prediction interval. Positive Hedges’ g favors extended-reality training.

**Table 2.** Study-level effects included in the primary meta-analysis.

| Study | XR<br>n | XR<br>mean<br>(SD) | Comparator<br>n | Comparator<br>mean (SD) | Outcome | Task | Comparator | Delay,<br>d | Hedges g<br>(95% CI) | Weight,<br>% | Source |
| --- | --- | --- | --- | --- | --- | --- | --- | --- | --- | --- | --- |
| Chou 2006 | 8 | 23.60<br>(5.40) | 8 | 22.90 (4.80) | OSATS quality<br>score (maximum<br>35) | Transfer | Active physical<br>model | 60 | 0.13<br>(−0.80 to<br>1.06) | 17.3 | PDF p. 4, Table 3; group<br>means/SDs directly reported |
| Sabench<br>Pereferrer<br>2013 | 24 | 96.57<br>(9.10) | 24 | 82.40 (13.85) | Fine-dissection<br>LapSim score (%) | Same | Inactive/no<br>training | 30 | 1.19 (0.58<br>to 1.80) | 17.7 | PDF p. 5, Table 3; three equal n=8<br>academic-year strata combined with<br>standard aggregate mean/SD<br>formulas |
| Jensen 2014 | 14 | 35.00<br>(6.90) | 14 | 29.60 (6.60) | Lobectomy<br>performance time<br>with penalty | Transfer | Active black-box trainer | 45 | −0.78<br>(−1.53 to<br>−0.03) | 17.6 | PDF p. 6, Table 1; penalized time<br>directly reported; lower-is-better<br>direction reversed |
| Nemani<br>2018 | 6 | 209.40<br>(21.00) | 5 | 155.00 (21.20) | VBLaST<br>normalized pattern-<br>cutting score | Same | Inactive/no<br>training | 14 | 2.36 (0.82<br>to 3.90) | 16.0 | PMC5809233 Results, Figure 4;<br>means/SDs directly reported in text |
| Cychosz<br>2018 | 22 | 30.09<br>(9.23) | 21 | 24.00 (10.13) | FAST composite<br>arthroscopy score | Transfer | Inactive/no<br>training | 11 | 0.62 (0.02<br>to 1.22) | 17.8 | PDF p. 5, Table 3; group<br>means/SDs directly reported |
| Linsk 2018 | 7 | 77.60<br>(3.80) | 9 | 44.70 (6.20) | VBLaST<br>normalized pattern-<br>cutting score | Same | Inactive/no<br>training | 14 | 5.86 (3.50<br>to 8.23) | 13.7 | PDF p. 7, Figure 3C; means/SDs<br>digitized; caption states that error<br>bars are SDs |

Positive Hedges' g favors extended-reality training. Weights are inverse-variance Paule-Mandel random-effects weights. Linsk 2018 values were digitized from

The six studies did not represent a single uniform training contrast. They differed in procedure, scale, same-task versus transfer assessment, and active versus inactive comparator. GRADE certainty for this broad comparative outcome was very low because of risk of bias, inconsistency, indirectness, and imprecision; publication bias could not be assessed reliably with six studies.

### Exploratory subgroup analyses

Same-task studies (k=3) produced a larger but very imprecise estimate (g=2.95, 95% CI −2.94 to 8.84) than transfer studies (k=3; g=0.01, 95% CI −1.79 to 1.81). Inactive-comparator studies (k=4) yielded g=2.30 (95% CI −1.27 to 5.88), whereas active-comparator studies (k=2) yielded g=−0.37 (95% CI −6.10 to 5.36). Short- and intermediate-window estimates were also imprecise. All three pooled same-task studies used inactive comparators, while two of three transfer studies used active comparators; task relation and comparator type were therefore confounded. These subgroup findings are descriptive rather than causal (Table 3).

**Table 3.**
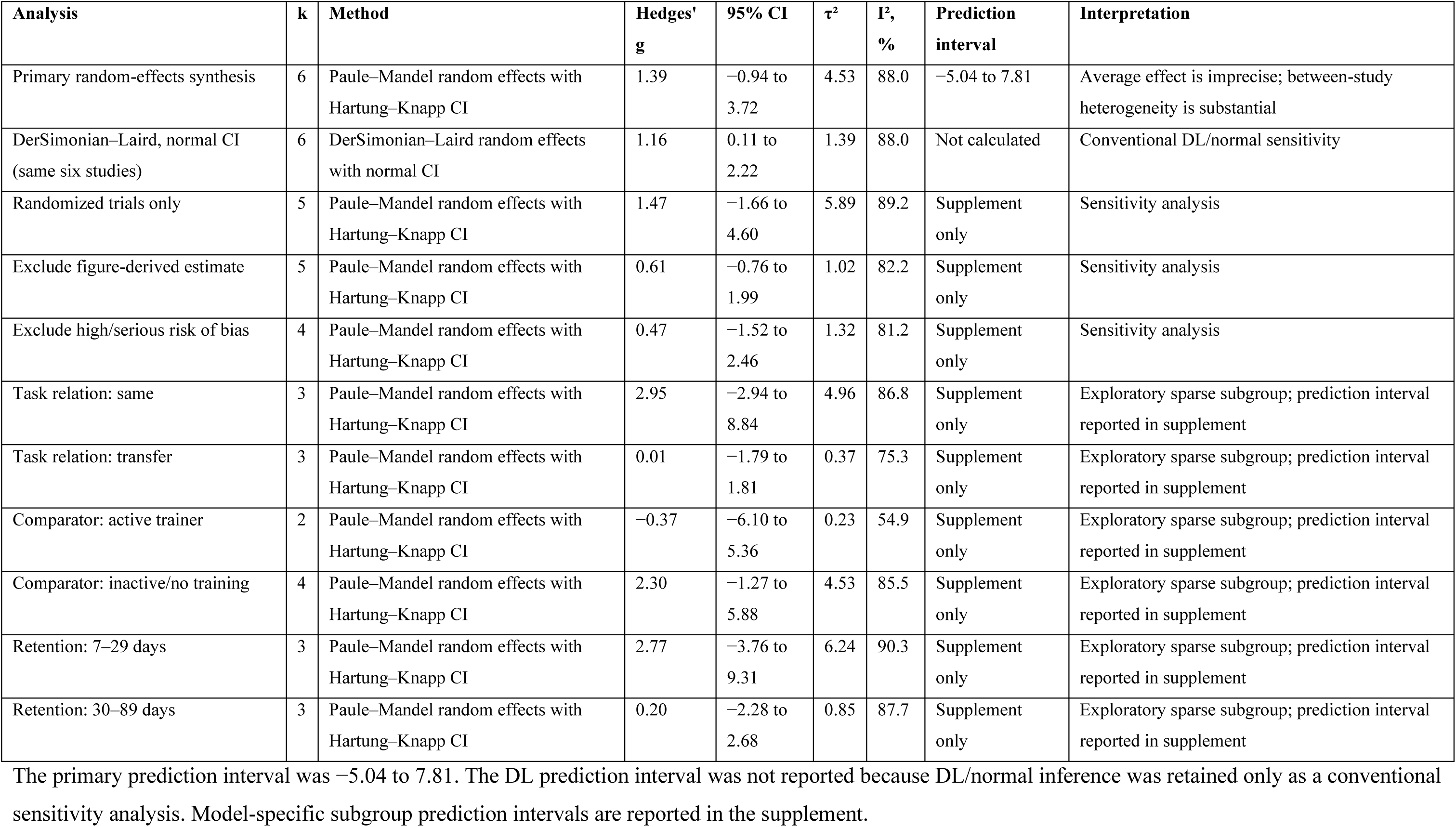
Meta-analysis, subgroup, and sensitivity results.

| Analysis | k | Method | Hedges' g | 95% CI | $\tau^2$ | I<sup>2</sup>, % | Prediction interval | Interpretation |
| --- | --- | --- | --- | --- | --- | --- | --- | --- |
| Primary random-effects synthesis | 6 | Paule–Mandel random effects with Hartung–Knapp CI | 1.39 | −0.94 to 3.72 | 4.53 | 88.0 | −5.04 to 7.81 | Average effect is imprecise; between-study heterogeneity is substantial |
| DerSimonian–Laird, normal CI (same six studies) | 6 | DerSimonian–Laird random effects with normal CI | 1.16 | 0.11 to 2.22 | 1.39 | 88.0 | Not calculated | Conventional DL/normal sensitivity |
| Randomized trials only | 5 | Paule–Mandel random effects with Hartung–Knapp CI | 1.47 | −1.66 to 4.60 | 5.89 | 89.2 | Supplement only | Sensitivity analysis |
| Exclude figure-derived estimate | 5 | Paule–Mandel random effects with Hartung–Knapp CI | 0.61 | −0.76 to 1.99 | 1.02 | 82.2 | Supplement only | Sensitivity analysis |
| Exclude high/serious risk of bias | 4 | Paule–Mandel random effects with Hartung–Knapp CI | 0.47 | −1.52 to 2.46 | 1.32 | 81.2 | Supplement only | Sensitivity analysis |
| Task relation: same | 3 | Paule–Mandel random effects with Hartung–Knapp CI | 2.95 | −2.94 to 8.84 | 4.96 | 86.8 | Supplement only | Exploratory sparse subgroup; prediction interval reported in supplement |
| Task relation: transfer | 3 | Paule–Mandel random effects with Hartung–Knapp CI | 0.01 | −1.79 to 1.81 | 0.37 | 75.3 | Supplement only | Exploratory sparse subgroup; prediction interval reported in supplement |
| Comparator: active trainer | 2 | Paule–Mandel random effects with Hartung–Knapp CI | −0.37 | −6.10 to 5.36 | 0.23 | 54.9 | Supplement only | Exploratory sparse subgroup; prediction interval reported in supplement |
| Comparator: inactive/no training | 4 | Paule–Mandel random effects with Hartung–Knapp CI | 2.30 | −1.27 to 5.88 | 4.53 | 85.5 | Supplement only | Exploratory sparse subgroup; prediction interval reported in supplement |
| Retention: 7–29 days | 3 | Paule–Mandel random effects with Hartung–Knapp CI | 2.77 | −3.76 to 9.31 | 6.24 | 90.3 | Supplement only | Exploratory sparse subgroup; prediction interval reported in supplement |
| Retention: 30–89 days | 3 | Paule–Mandel random effects with Hartung–Knapp CI | 0.20 | −2.28 to 2.68 | 0.85 | 87.7 | Supplement only | Exploratory sparse subgroup; prediction interval reported in supplement |

The primary prediction interval was −5.04 to 7.81. The DL prediction interval was not reported because DL/normal inference was retained only as a conventional sensitivity analysis. Model-specific subgroup prediction intervals are reported in the supplement.

### Sensitivity analyses

Using DerSimonian-Laird estimation with a normal confidence interval gave g=1.16 (95% CI 0.11 to 2.22), which was more apparently precise than the primary small-sample-robust analysis. Restriction to randomized trials gave g=1.47 (95% CI −1.66 to 4.60). Excluding the figure-derived estimate reduced the summary to g=0.61 (95% CI −0.76 to 1.99); excluding studies at high or serious risk of bias gave g=0.47 (95% CI −1.52 to 2.46). Every leave-one-out Hartung-Knapp interval crossed the null. Across these analyses, changes in point estimates and interval width reinforced that the evidence was too sparse and heterogeneous for a stable single estimate.

### Narrative findings from nonpooled studies

Most studies could not be pooled because they lacked an independent delayed comparator, reported incompatible outcomes, supplied median-based summaries without an appropriate variance estimate, or displayed values graphically without defined error bars. Others compared XR strategies or included a booster or retraining exposure before the delayed endpoint. Exclusion from the meta-analysis therefore reflected incompatibility with the prespecified contrast, not absence of delayed information. This distinction retained relevant delayed evidence while preventing designs with different educational questions from being combined into a misleading single quantitative estimate overall.

Nonpooled pure-retention studies provided same-task outcomes more often than clinically proximal transfer. Several short- or intermediate-term reports described persistence of some trained performance, but scoring differences, small groups, and incomplete dispersion data prevented a common estimate. Transfer was tested across dissimilar physical trainers, anatomical models, tissue tasks, and simulator systems. Similar performance against an active trainer addressed comparative effectiveness rather than whether either group had learned.

Longer-term evidence depended largely on small uncontrolled cohorts. In one cohort, median performance at six months remained above baseline despite an uncertain reduction from immediate performance; by 18 months, it had returned to baseline [21]. Other studies assessed approximately three to seven months, but clinical exposure, simulator access, acclimatization attempts, attrition, or lack of a concurrent control limited attribution to the original XR course [15,19,29,30,40,44–46]. These studies did not establish a common decay rate.

Maintenance and reacquisition findings were complementary. One randomized experiment found that a scheduled practice session attenuated short-term deterioration compared with no interim practice [23]. Other maintenance designs incorporated repeated practice and therefore evaluated preservation under continuing exposure [42,45]. Some small studies reported faster reacquisition than initial acquisition [16,41]. This may be educationally useful but does not show that performance remained intact during the interval.

## Discussion

This review found a broad but methodologically uneven literature on delayed technical performance after XR surgical simulation. Although 33 studies were included, only six provided an independent, uncontaminated delayed contrast with compatible data. The primary estimate was positive, but its wide confidence interval, substantial heterogeneity, and prediction interval across the null indicate that delayed benefit varies by task, comparator, curriculum, measurement, and context rather than representing a single durable XR effect.

The pooled point estimate was large in magnitude, but the Hartung-Knapp confidence interval (-0.94 to 3.72), substantial heterogeneity, and broad prediction interval (-5.04 to 7.81) show that the average remains highly uncertain. The estimate is compatible with effects favoring XR, effects near the null, and effects favoring the comparator. This very low-certainty result is therefore not a stable estimate of a common durable benefit.

The prediction interval addresses dispersion of underlying effects in comparable future settings, rather than uncertainty around the average alone. Its breadth indicates that the pooled mean is not reliably transportable across procedures, curricula, tasks, or comparator intensities. It is not a forecast for an individual learner, and its extremes should not be read as literal clinical magnitudes.

The Linsk study supplied the largest effect and was digitized from Figure 3C on PDF page 7 [37]. The source-resolution image was calibrated to the figure’s linear 0-90 axis using all ten horizontal gridlines in Pillow version 12.3.0. The retention means and standard deviations were independently verified by a second reviewer against the figure and caption, which defines the error bars as standard deviations; calibration coordinates and extracted values are archived with the supplement. Excluding this small, high-risk estimate reduced the summary from g = 1.39 to 0.61 (95% CI -0.76 to 1.99). Every leave-one-out Hartung-Knapp interval crossed the null, confirming that the pooled magnitude is sensitive to individual studies.

**Figure 3.**
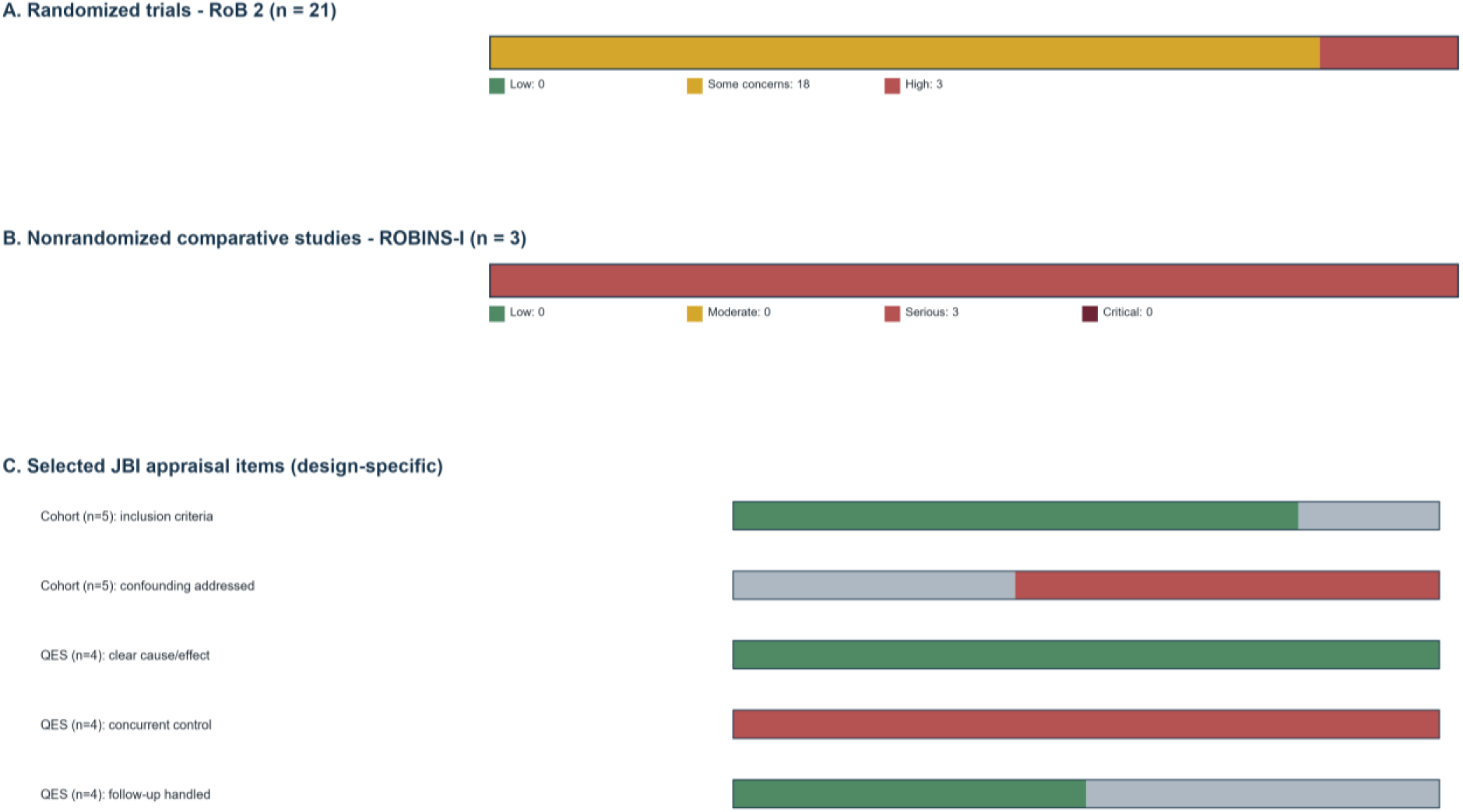
Risk-of-bias summary by study design. RoB 2 and ROBINS-I global judgments are shown on their respective scales. The JBI panel shows selected, design-specific cohort and quasi-experimental checklist items; complete item-level appraisals are in the supplement, and no global JBI score was assigned.

These findings complement broader evidence that simulation can improve technical performance and transfer in selected settings [1–3,6–10], while procedural-skill decay varies across tasks, learners, and intervals [4,5]. A small cohort in this review remained above baseline at six months but returned to baseline by 18 months [21], illustrating that residual learning can coexist with loss from peak performance without establishing a universal decay trajectory.

Same-task persistence appeared stronger than transfer, but the subgroup estimates arose from only three studies each and were entangled with comparator type. Repetition of the trained platform preserves the perceptual cues, instrument behavior, and scoring structure encountered during practice. It may therefore reveal a durable task representation while also benefiting from device familiarity. Transfer requires learners to identify which components of that representation remain useful when tissue properties, visual cues, instrument dynamics, or procedural context change. This is a harder test and often a more clinically relevant one. A weaker transfer estimate does not mean that same-task learning is unimportant, but it limits claims that proficiency on the simulator will persist as improved performance elsewhere. Delayed studies should ideally measure both outcomes so specificity and generalization can be examined within the same participants and interval.

Comparator choice further complicates interpretation. An inactive control asks whether XR practice is better than no technical practice. A box trainer, physical model, or alternative simulation curriculum asks whether XR adds value over another form of deliberate practice. The latter is a stricter and often more useful procurement or curriculum question. Two of the three pooled transfer studies used active comparators, while all pooled same-task studies used inactive controls. A near-zero effect against an effective physical trainer should therefore not be interpreted as evidence that XR fails to teach; it may indicate that two training routes produce similar delayed performance. Conversely, a large effect against no practice does not establish superiority over a lower-cost active method. Future syntheses will need enough studies to separate task relation from comparator type rather than treating the observed subgroup contrast as causal.

Pure retention, maintenance, and reacquisition inform different curricular decisions. Pure retention concerns performance without targeted practice; maintenance evaluates whether planned exposure prevents deterioration; and reacquisition measures the effort required to restore performance. A scheduled session attenuated short-term deterioration in one randomized experiment [23], and some studies reported faster reacquisition than initial learning [16,41], but neither finding defines a universal refresher interval. Reassessment should reflect procedure-specific risk, frequency, prior expertise, and the required proficiency standard. Programs should record end-of-training performance, intervening exposure, delayed performance before refreshers, and the practice needed to regain criterion.

Strengths include the seven-day minimum, explicit separation of same-task retention from transfer, distinction among pure retention, maintenance, and reacquisition, linkage of companion reports, and one endpoint per independent study family. The analysis used small-sample-robust random-effects inference, a transparent eligibility map, design-appropriate risk-of-bias tools, and an explicit GRADE assessment, while retaining relevant nonpooled evidence in narrative synthesis.

Limitations include six pooled studies and 162 selected-arm participants, substantial heterogeneity, and no pooled follow-up of at least 90 days. Samples were small, delayed attrition was common, allocation methods were often incompletely reported, and outcomes varied across speed, errors, procedural quality, and composites. Some reports lacked usable variance estimates, clinically proximal transfer was sparse, and intervening practice was not always controlled. The broad estimand combined clinically different contrasts, correlated design features prevented causal subgroup interpretation, the mean was influenced by a figure-derived estimate, and publication bias could not be assessed reliably.

Because standardized effects combined different technical-performance measures, the pooled Hedges’ g should not be interpreted as a common clinically meaningful difference across procedures. Future trials should predeclare an objective delayed endpoint assessed before refresher exposure and report arm-specific sample sizes, means, standard deviations, and attrition. Training dose, end-of-training proficiency, interim clinical cases, and simulation access should be documented. Validated quality measures should accompany speed metrics, and the same cohort should undergo both trained-task and clinically meaningful transfer assessments when feasible. Active comparators, multicenter recruitment, follow-up beyond three months, randomized booster schedules, and standardized reacquisition-to-criterion measures would improve future synthesis.

## Conclusions

Delayed technical advantages after extended-reality simulation were observed in some settings, but the comparative evidence does not define a precise or generalizable retention effect. The six-study average remained uncertain under small-sample-robust inference and was accompanied by substantial heterogeneity. Effects were larger against inactive than active comparators, although task relation and comparator type were confounded. Transfer evidence was sparse, and no pooled study assessed an interval of 90 days or longer.

## Declarations

### Protocol and registration

This systematic review was not prospectively registered on PROSPERO; however, the study protocol was fully established and finalized prior to the commencement of data extraction and analysis.

### Ethics approval

Not applicable; this study synthesized published data.

### Data availability

The data supporting this synthesis are available from the corresponding author on reasonable request.

