## Supplementary files for "Delayed Retention of Technical Surgical Skills Following Virtual, Augmented, and Mixed-Reality Simulation: A Systematic Review and Meta-analysis"

#### Table S1. Electronic search strategies

| Source | Variant | Platform | Coverage | Exact strategy | Filters/limits | Records identified | Yield basis |
|---|---|---|---|---|---|---|---|
| PubMed/MEDLINE | Broad main search | PubMed (National Library of Medicine) | Inception to 2026-05-05 | ("Virtual Reality"[Mesh] OR "Augmented Reality"[Mesh] OR "virtual reality"[tiab] OR "augmented reality"[tiab] OR "mixed reality"[tiab] OR "extended reality"[tiab] OR "immersive virtual reality"[tiab] OR "virtual simulation"[tiab] OR "virtual simulator*"[tiab] OR "immersive simulation"[tiab]) AND ("Simulation Training"[Mesh] OR simulat*[tiab] OR train*[tiab] OR educat*[tiab] OR learn*[tiab]) AND ("Surgical Procedures, Operative"[Mesh] OR surg*[tiab] OR laparoscop*[tiab] OR robotic surg*[tiab] OR microsurg*[tiab] OR endoscop*[tiab] OR arthroscop*[tiab] OR operative[tiab]) AND ("Clinical Competence"[Mesh] OR "Psychomotor Performance"[Mesh] OR skill*[tiab] OR competenc*[tiab] OR proficien*[tiab] OR performance[tiab] OR dexterity[tiab] OR accuracy[tiab] OR error*[tiab] OR technical[tiab]) | Publication date from database inception through 2026-05-05; no language or full-text filter | 3990 | Official PubMed E-utilities reconstruction |
| PubMed/MEDLINE | Retention safety-net search | PubMed (National Library of Medicine) | Inception to 2026-05-05 | ("virtual reality"[tiab] OR "augmented reality"[tiab] OR "mixed reality"[tiab] OR "extended reality"[tiab] OR "virtual simulation"[tiab] OR "virtual simulator*"[tiab]) AND (surg*[tiab] OR laparoscop*[tiab] OR robotic*[tiab] OR microsurg*[tiab] OR endoscop*[tiab]) AND (retention[tiab] OR retained[tiab] OR "skill retention"[tiab] OR "skill decay"[tiab] OR "skills decay"[tiab] OR "skill maintenance"[tiab] OR maintenance[tiab] OR persistence[tiab] OR sustained[tiab] OR "long term"[tiab] OR "long-term"[tiab] OR longitudinal[tiab] OR follow-up[tiab] OR followup[tiab] OR delayed[tiab] OR retest*[tiab] OR "transfer of training"[tiab] OR transferability[tiab]) | Publication date from database inception through 2026-05-05; no language or full-text filter | 905 | Official PubMed E-utilities reconstruction |
| Europe PMC | Broad main search | Europe PMC | Inception to 2026-05-05 | (TITLE_ABS:"virtual reality" OR TITLE_ABS:"augmented reality" OR TITLE_ABS:"mixed reality" OR TITLE_ABS:"extended reality" OR TITLE_ABS:"immersive virtual reality" OR TITLE_ABS:"virtual simulation" OR TITLE_ABS:"virtual simulator" OR TITLE_ABS:"immersive simulation") AND (TITLE_ABS:surgery OR TITLE_ABS:surgical OR TITLE_ABS:laparoscopic OR TITLE_ABS:laparoscopy OR TITLE_ABS:robotic OR TITLE_ABS:microsurgery OR TITLE_ABS:endoscopic OR TITLE_ABS:arthroscopic) AND (TITLE_ABS:simulation OR TITLE_ABS:training OR TITLE_ABS:education OR TITLE_ABS:learning) AND (TITLE_ABS:skill OR TITLE_ABS:skills OR TITLE_ABS:competence OR TITLE_ABS:proficiency OR TITLE_ABS:performance OR TITLE_ABS:dexterity OR TITLE_ABS:accuracy OR TITLE_ABS:technical) | FIRST_PDATE from database inception through 2026-05-05; no language or full-text filter | 2881 | Official Europe PMC REST API reconstruction |
| Europe PMC | Retention safety-net search | Europe PMC | Inception to 2026-05-05 | (TITLE_ABS:"virtual reality" OR TITLE_ABS:"augmented reality" OR TITLE_ABS:"mixed reality" OR TITLE_ABS:"extended reality" OR TITLE_ABS:"virtual simulation") AND (TITLE_ABS:surgery OR TITLE_ABS:surgical OR TITLE_ABS:laparoscopic OR TITLE_ABS:robotic OR TITLE_ABS:microsurgery) AND (TITLE_ABS:retention OR TITLE_ABS:"skill retention" OR TITLE_ABS:"skill decay" OR TITLE_ABS:"skill maintenance" OR TITLE_ABS:longitudinal OR TITLE_ABS:"follow-up" OR TITLE_ABS:delayed OR TITLE_ABS:retest OR TITLE_ABS:"transfer of training") | FIRST_PDATE from database inception through 2026-05-05; no language or full-text filter | 426 | Official Europe PMC REST API reconstruction |
| CENTRAL | Broad main search | Cochrane Library, CENTRAL (2026, Issue 5) | Database coverage through 2026-05-05 | ("virtual reality" OR "augmented reality" OR "mixed reality" OR "extended reality" OR "immersive virtual reality" OR "virtual simulation" OR "virtual simulator" OR "virtual simulators" OR "immersive simulation") AND (surg* OR laparoscop* OR robotic* OR microsurg* OR endoscop* OR arthroscop* OR operative) AND (simulat* OR train* OR educat* OR learn*) AND (skill* OR competen* OR proficien* OR performance OR dexterity OR accuracy OR error* OR technical OR psychomotor) | CENTRAL trials; coverage through 2026-05-05; no language filter | Reported in combined CENTRAL source total below | CENTRAL results reported as one source-level total |
| CENTRAL | Retention safety-net | Cochrane Library, CENTRAL (2026, | Database coverage | ("virtual reality" OR "augmented reality" OR "mixed reality" OR "extended reality" OR "immersive virtual reality" OR "virtual | CENTRAL trials; coverage through | Reported in combined | CENTRAL results reported as one |

| | | | | | | | |
|---|---|---|---|---|---|---|---|
| | search | Issue 5) | through 2026-05-05 | simulation" OR "virtual simulator" OR "virtual simulators" OR "immersive simulation") AND (surg* OR laparoscop* OR robotic* OR microsurg* OR endoscop* OR arthroscop* OR operative) AND (retention OR retained OR "skill retention" OR "skill decay" OR "skills decay" OR maintenance OR persistence OR sustained OR longitudinal OR "follow-up" OR followup OR delayed OR retest* OR "transfer of training") | 2026-05-05; no language filter | CENTRAL source total below | source-level total |
| CENTRAL | Combined source total | Cochrane Library, CENTRAL (2026, Issue 5) | Database coverage through 2026-05-05 | Broad main plus retention safety-net variants shown above | Source-level aggregate used in the PRISMA record total | 115 | Source-level reconciliation within the 8,628-record aggregate |
| ERIC | Broad main search | ERIC API (Institute of Education Sciences) | Inception to 2026-05-05 | ("virtual reality" OR "augmented reality" OR "mixed reality" OR "extended reality") AND (surgery OR surgical OR laparoscopic OR robotic OR microsurgery OR endoscopic) AND (simulation OR training OR education OR learning) AND (skill OR skills OR performance OR proficiency OR competence OR technical) | Publication date through 2026-05-05; no language or full-text filter | 24 | Official ERIC API reconstruction |
| ERIC | Retention safety-net search | ERIC API (Institute of Education Sciences) | Inception to 2026-05-05 | ("virtual reality" OR "augmented reality" OR "mixed reality" OR "extended reality") AND (surgery OR surgical OR laparoscopic OR robotic OR microsurgery) AND (retention OR "skill retention" OR "skill decay" OR longitudinal OR "follow-up" OR delayed OR retest) | Publication date through 2026-05-05; no language or full-text filter | 2 | Official ERIC API reconstruction |
| ClinicalTrials.gov | General surgical simulation search | ClinicalTrials.gov API v2 | Records first posted through 2026-05-05 | ("virtual reality" OR "augmented reality" OR "mixed reality") AND (surgical OR surgery) AND (simulation OR training) | Study First Posted on or before 2026-05-05; all recruitment statuses; no country or phase filter | 205 | Official ClinicalTrials.gov API v2 reconstruction |
| ClinicalTrials.gov | Procedure-specific safety-net search | ClinicalTrials.gov API v2 | Records first posted through 2026-05-05 | ("virtual reality" OR "augmented reality" OR "mixed reality") AND (laparoscopy OR laparoscopic OR robotic surgery OR microsurgery) | Study First Posted on or before 2026-05-05; all recruitment statuses; no country or phase filter | 80 | Official ClinicalTrials.gov API v2 reconstruction |

The 8,628-record identification total is reproduced by the source/variant yields in Table S1. The PubMed, Europe PMC, ERIC, and ClinicalTrials.gov figures are retrospective reconstructions using the documented strings and May 5, 2026 ceiling; the two CENTRAL variants are available only as a combined source total of 115. The original contemporaneous export and separate CENTRAL variant yields require author confirmation.

#### Table S2. Operational definitions

| Term | Definition |
|---|---|
| Pure retention | Objective delayed performance measured before any relevant task-specific booster, refresher, or retraining exposure. |
| Primary endpoint selection | First eligible uncontaminated delayed assessment when multiple retention assessments were reported, unless the prespecified endpoint rule indicated otherwise. |
| Maintenance | Delayed objective performance after planned intermittent or refresher practice. |
| Reacquisition | Performance after retraining begins following a delay. |
| Same task | Delayed assessment on the trained simulator and task. |
| Transfer | Delayed assessment on another simulator, physical model, cadaver, animal model, or operative task. |
| Short follow-up | 7–29 days |
| Intermediate follow-up | 30–89 days |
| Longer-term follow-up | At least 90 days |

#### Table S3. Cychosz companion-report study family

| Report | Citation | Study-family linkage | Review role |
|---|---|---|---|
| Cychosz et al. 2018 | Arthroscopy 2018;34(5):1543–1549 | Same 43-participant cohort | Primary report supplying the quantitative contrast |
| Cychosz et al. 2019 | Iowa Orthop J. 2019;39(1):7–13 | Same 43-participant cohort | Companion report informing study context; not an additional study or meta-analysis unit |

### Table S4. Detailed included-study characteristics

| Study ID | Authors | Title | Year | Journal | DOI | Country | Design | Population and sample | XR intervention | Comparator | Delayed assessment | Retention interval | Retention window | Task relation | Pure retention | Primary meta-analysis | Review disposition | Risk-of-bias tool | Risk-of-bias judgment | Risk-of-bias evidence |
|---|---|---|---|---|---|---|---|---|---|---|---|---|---|---|---|---|---|---|---|---|
| STUDY_001 | Kenneth Ogan; Lucas Jacomides; Michael J. Shulman; Claus G. Roehrborn; Jeffrey A. Cadeddu; Margaret S. Pearle | Virtual ureteroscopy predicts ureteroscopic proficiency of medical students on a cadaver | 2004 | Journal of Urology | 10.1097/01.ju.0000131631.60022.d9 | United States | Prospective cohort with trained students and resident expertise reference | 32 total: 16 medical students and 16 urology residents (PDF p1) | Students: 10 supervised 30-min sessions (5 h) on Uromentor VR over 2 weeks; baseline and repeat same VR case | Untrained residents (no VR training) as expertise/reference group; cadaver comparison is not a causal untrained comparator | 2 weeks after baseline (VR2) | 14 days | 7-29 days | SAME + TRANSFER | YES | NO | Systematic review, nonpooled | JBI cohort checklist (condensed items) | No global score | Methods and Tables 2/4 (PDF pp. 1-3): trained students with resident reference; objective simulator and cadaver ratings; no causal untrained comparator. |
| STUDY_002 | Dimitrios Stefanidis; James R. Korndorffer Jr; Rafael Sierra; Cheri Touchard; J. Bruce Dunne; Daniel J. Scott | Skill retention following proficiency-based laparoscopic simulator training | 2005 | Surgery | 10.1016/j.surg.2005.06.002 | United States | Prospective within-cohort retention study | 14 surgery residents R1-R4 with no previous/minimal VR or VT experience; six R1 four R2 two R3 two R4 (PDF pp1; 3) | Proficiency-based curriculum: 12 MIST-VR and 5 videotrainer tasks to criterion on 2 consecutive repetitions | No concurrent control; 3 experts tested only as proficiency benchmark | Posttest 13.2 +/- 11.8 days after proficiency; retention 7.0 +/- 4.0 months after proficiency (PDF pp1; 3) | ~7 months | >=90 days | SAME | YES | NO | Systematic review, nonpooled | JBI cohort checklist (condensed items) | No global score | Methods/Results (PDF pp. 1-3): convenience cohort of 14 residents; automated MIST-VR outcomes; all completed delayed assessment. |
| STUDY_003 | J. A. Windsor; F. Zoha | The laparoscopic performance of novice surgical trainees: testing for acquisition, loss, and reacquisition of psychomotor skills | 2005 | Surgical Endoscopy | 10.1007/s00464-004-2200-9 | New Zealand | Two-session prospective repeated-measures cohort | 10 junior surgical registrars/novice surgical trainees with little/no prior laparoscopic exposure (PDF pp1-2); 10 experienced surgeons define criterion level | Two identical MIST-VR training sessions one month apart on stretch diathermy (SD) and manipulation diathermy (MD) | Experienced surgeons are criterion/reference participants | not an intervention comparator | ~1 month | 30-89 days / borderline depending exact day | SAME | PARTLY | NO | Systematic review, nonpooled | JBI cohort checklist (condensed items) | No global score | Methods and Tables 4-5 (PDF pp. 1-6): repeated-measures cohort with expert reference only; delayed retention and reacquisition reported. |
| STUDY_004 | David S. Chou; Corollos Abdelshehid; Ralph V. Clayman; Elspeth M. McDougall | Comparison of results of virtual-reality simulator and training model for basic ureteroscopy training | 2006 | Journal of Endourology | | United States | Randomized controlled trial | 16 first-year medical students with no prior endourology or VRS experience (PDF pp1-2) | Up to 2 h supervised training on URO-Scopic materials ureteroscopy model (TMU) or Simbionix UROMentor VRS | TMU versus VRS randomized groups (8 per group; group labels in methods/results) | 2 months after training (PDF pp1 | 2 months | 30-89 days | TRANSFER | YES | YES | Primary meta-analysis | RoB 2 | Some concerns | Methods/Table 3 (PDF pp. 1-4): randomization stated without concealment details; all 16 assessed; OSATS rated by one unblinded observer. |
| STUDY_005 | E. G. G. Verdaasdonk; L. P. S. | The influence of different | 2007 | Surgical Endoscopy | 10.1007/s00464-005-0852-8 | Netherlands | Randomized controlled trial | 20 endoscopy-naive | 12 repetitions of three SIMENDO VR tasks (drop | Group B same VR training dose | Posttest 7 days after last training session | 7 days | 7-29 days | SAME | YES | NO | Systematic review, | RoB 2 | Some concerns | Methods/Table 2/Figure 4 (PDF pp. 1-5): |

| | | | | | | | | | | | | | | | | | | | | |
|---|---|---|---|---|---|---|---|---|---|---|---|---|---|---|---|---|---|---|---|---|
| | Stassen; R. P. J. van Wijk; J. Dankelman | training schedules on the learning of psychomotor skills for endoscopic surgery | | | | | | students randomized: distributed over 3 days group A n=10 vs distributed within 1 day group B n=10 (PDF pp1-2) | balls; ring/needle manipulation; 30-degree endoscope) | in one day with 15-min breaks; group A over 3 consecutive days | (PDF pp1-2) | | | | | | nonpooled | | | randomized n=20; complete follow-up and automated metrics; concealment/protocol unavailable. |
| STUDY_006 | Christopher W. Snyder; Marianne J. Vandromme; Sharon L. Tyra; Mary T. Hawn | Retention of colonoscopy skills after virtual reality simulator training by independent and proctored methods | 2010 | The American Surgeon | | United States | Randomized follow-up cohort from prior trial | 13 medical students at follow-up: 8 proctored and 5 independent (original cohort 32; PDF pp1-2) | 8-week proficiency-based AccuTouch Lower GI VR colonoscopy training with automated feedback; proctored arm adds expert feedback | Independent automated feedback versus proctored human plus simulator feedback | Median 4.5 months without simulator practice (ranges by group in Table 2 p3); baseline and posttraining also assessed | Median ~4.5 months | Cannot assign all participants reliably to 30-89 vs >=90 | SAME | YES | NO | Systematic review, nonpooled | ROBINS-I | Serious | Methods/Table 2 (PDF pp. 1-3): only 13/32 parent-study participants returned; follow-up selection was substantial and imbalanced. |
| STUDY_007 | Irina Kruglikova; Teodor P. Grantcharov; Asbjorn M. Drewes; Peter Funch-Jensen | The impact of constructive feedback on training in gastrointestinal endoscopy using high-fidelity virtual-reality simulation: a randomised controlled trial | 2009 | Gut | 10.1136/gut.2009.191825 | Russian Federation | Randomized controlled trial | 22 inexperienced trainees plus 8 expert colonoscopists (PDF p1); one feedback trainee had 10 rather than 15 repetitions and one non-feedback trainee missed follow-up (PDF p2) | 15 repetitions of AccuTouch Endoscopy simulator task 3 over 4 sessions/1 month; structured concurrent expert feedback vs simulator feedback | Non-feedback control receives virtual attending/simulator feedback only | Delayed simulator retention task 3 and near-transfer task 5 at 4-6 weeks after last repetition (PDF p2) | 4-6 weeks | Crosses 7-29 and 30-89 day windows | SAME / near-transfer | YES | NO | Systematic review, nonpooled | RoB 2 | Some concerns | Methods/Figure 1 (PDF pp. 1-4): sealed-envelope allocation; one training deviation and one missed delayed test; automated metrics. |
| STUDY_008 | Mathilde Maagaard; Jette Led Sorensen; Jeanett Oestergaard; Torur Dalsgaard; Teodor P. Grantcharov; Bent S. Ottesen; Christian Rifbjerg Larsen | Retention of laparoscopic procedural skills acquired on a virtual-reality surgical trainer | 2010 | Surgical Endoscopy | 10.1007/s00464-010-1233-5 | Denmark | Longitudinal follow-up cohort | Novices <5 procedures n=9 (trainees) and experts >200 procedures in prior 3 years n=10; novice follow-up n=8 at 18 months after one dropout (PDF pp1-4) | LapSim VR ectopic pregnancy/salpingectomy module; initial training to proficiency | 10 sessions | Expert group is expertise reference and continued clinical laparoscopy; no randomized untrained comparator | 6 and 18 months | >=90 days | SAME | YES | NO | Systematic review, nonpooled | JBI cohort checklist (condensed items) | No global score | Methods/Tables 1-2 (PDF pp. 1-4): small novice cohort with expert reference; one novice lost by 6 months and another by 18 months. |
| STUDY_009 | Mubashir Mulla; Davendra Sharma; Masood Moghul; | Learning basic laparoscopic skills: a randomized controlled | 2012 | Journal of Surgical Education | 10.1016/j.jsurg.2011.07.011 | United Kingdom | Randomized controlled trial | 41 medical students with no prior laparoscopic | Training varied by group; VRS group received induction and practice on CAE Healthcare VR | Control no extra training; all groups assessed on VRS and box trainer | 1 week after initial demonstration/training (PDF pp1-3) | 1 week | 7-29 days | SAME + TRANSFER | YES | NO | Systematic review, nonpooled | RoB 2 | Some concerns | Methods/Tables 1-2 (PDF pp. 1-3): randomized allocation reported; arm-level delayed |

| | | | | | | | | | | | | | | | | | | | | |
|---|---|---|---|---|---|---|---|---|---|---|---|---|---|---|---|---|---|---|---|---|
| | Obeda Kailani; Judith Dockery; Salma Ayis; Philippe Grange | study comparing box trainer, virtual reality simulator, and mental training | | | | | | experience randomized to 5 groups (control; box; enhanced box; VRS; mental; PDF pp1-2) | lap simulator; box groups received pelvic/box trainer; mental group mental practice | | | | | | | | | | | denominators incompletely reported; objective simulator/box-trainer metrics. |
| STUDY_010 | Anthony G. Gallagher; Julie Anne Jordan-Black; Gerald C. O'Sullivan | Prospective, randomized assessment of the acquisition, maintenance, and loss of laparoscopic skills | 2012 | Annals of Surgery | 10.1097/SLA.0b013e318251f3d2 | Ireland | Two prospective randomized studies | Study 1: 24 laparoscopic novices; Study 2: 16 novices (12 male) with no previous laparoscopic surgery (PDF pp1-2 | 4) | Study 2 all trained identical MIST-VR curriculum; Practice arm received one complete simulator trial 1 week later | No-practice arm received no intervening trial; both assessed 2 weeks after training | 2 weeks | 7-29 days | SAME | NO | NO | Systematic review, nonpooled | RoB 2 | Some concerns | Study 2 Methods/Results and Figures 4-6 (PDF pp. 4-5): randomized booster/no-booster experiment; n=16 analyzed; automated outcomes. |
| STUDY_011 | Zhang L; Sankaranarayanan G; Arikatla VS; Ahn W; Grosdemouge C; Rideout JM; Epstein SK; De S; Schwaitzberg SD; Jones DB; Cao CGL | Characterizing the learning curve of the VBLaST-PT (Virtual Basic Laparoscopic Skill Trainer) | 2013 | Surgical Endoscopy | 10.1007/s00464-013-2932-5 | United States | randomized 3-arm trial | 18 medical students (6 VBLaST; 6 FLS; 6 control) | 150 peg-transfer trials over 3 weeks, 5 days/week on VBLaST-PT | FLS training and no-training control; delayed cross-system test | 2 weeks after post-test | 14 days | 7-29 days | SAME (primary); transfer possible separately | YES | NO | Systematic review, nonpooled | RoB 2 | Some concerns | Methods (PDF pp. 1-4): 18 students randomized 6/group; completion of all training trials varied; FLS outcomes manually video-scored. |
| STUDY_012 | Janse JA; Goedegebuure RSA; Veersema S; Broekmans FJM; Schreuder HWR | Hysteroscopic Sterilization Using a Virtual Reality Simulator: Assessment of Learning Curve | 2013 | Journal of Minimally Invasive Gynecology | 10.1016/j.jmig.2013.04.016 | Netherlands | prospective multicenter repeated-measures study | 30 novice medical students and 5 expert gynecologists | 9 bilateral Essure repetitions on HystSim/EssureSim; second series after delay | Expert gynecologists are reference/validation group | median 14 days (range 7-21) | Median 14 days (range 7-21) | 7-29 days | SAME | YES | NO | Systematic review, nonpooled | JBI cohort checklist (condensed items) | No global score | Methods/Table 3 (PDF pp. 1-6): 30 novices with expert reference; repeated objective assessments; delayed follow-up completeness not fully clear. |
| STUDY_013 | Sabench Pereferrer F; Hernández González M; Muñoz García A; Cabrera Vilanova A; Del Castillo Déjardin D | Evaluation of Surgical Skills in Medical Students Using a Virtual Simulator | 2013 | Cirugía Española | 10.1016/j.ciresp.2012.05.019 | Spain | prospective controlled training study | 48 medical students: 24 training (8 each 2nd/4th/6th year) and 24 controls | 3 sessions over 3 weeks on LapSim with 7 basic modular exercises | year-matched no-training controls | 30 days after final training/washout | 30 days | 30-89 days | SAME | YES | YES | Primary meta-analysis | ROBINS-I | Serious | Methods/Table 3 (PDF pp. 1-5): academic-year strata allocated to training/control without clear randomization; confounding by year and experience. |
| STUDY_014 | Jensen K; Ringsted C; Hansen HJ; Petersen RH; Konge L | Simulation-based training for thoracoscopic lobectomy: a randomized controlled trial: virtual-reality | 2014 | Surgical Endoscopy | 10.1007/s00464-013-3392-7 | Denmark | randomized controlled trial | 28 surgical residents (14 VR; 14 black-box) | VR nephrectomy module training versus traditional black-box simulator training | black-box simulation | VR mean 33.4±7.5 days; black-box 56.7±6.6 days | ~33-57 days | 30-89 days | TRANSFER | YES | YES | Primary meta-analysis | RoB 2 | Some concerns | Methods/Table 1 (PDF pp. 1-7): randomized residents; two VR dropouts before testing; objective time/error assessment; protocol unavailable. |

| | | | | | | | | | | | | | | | | | | | | |
|---|---|---|---|---|---|---|---|---|---|---|---|---|---|---|---|---|---|---|---|---|
| | | versus black-box simulation | | | | | | | | | | | | | | | | | | |
| STUDY_015 | Bjerrum F; Maagaard M; Sorensen JL; Larsen CR; Ringsted C; Winkel P; Ottesen B; Strandbygaard J | Effect of Instructor Feedback on Skills Retention After Laparoscopic Simulator Training: Follow-Up of a Randomized Trial | 2015 | Journal of Surgical Education | 10.1016/j.jsurg.2014.06.013 | Denmark | 6-month follow-up of randomized trial | 99 surgical novices initially; 65 completed follow-up | proficiency-based LapSim salpingectomy with/without instructor feedback | feedback vs no-feedback; both retrained to proficiency at follow-up | 182-262 days (intervention) and 198-? days (control); initial-to-follow-up median 214 days | 6 months | >=90 days | SAME | NO | NO | Systematic review, nonpooled | RoB 2 | High | Methods/Figure 1/Results (PDF pp. 1-5): 65/99 returned at 6 months and retrained to proficiency; substantial missing delayed data. |
| STUDY_016 | Van Bruwaene S; Schijven MP; Napolitano D; De Win G; Miserez M | Porcine Cadaver Organ or Virtual-Reality Simulation Training for Laparoscopic Cholecystectomy: A Randomized, Controlled Trial | 2015 | Journal of Surgical Education | 10.1016/j.jsurg.2014.11.015 | Belgium | prospective randomized 3-arm trial | 30 participants (10 control; 10 porcine organ; 10 VR) | basic laparoscopic training then VR LapMentor cholecystectomy proficiency training | control no additional training and porcine cadaver organ training | 4 months after training | 4 months | >=90 days | TRANSFER | YES | NO | Systematic review, nonpooled | RoB 2 | Some concerns | Methods/Tables 2-3 (PDF pp. 1-4): randomized 10/group; complete delayed assessment; quality ratings and concealment details limited. |
| STUDY_017 | Andersen, Steven Arild Wuyts | Retention of Mastoidectomy Skills After Virtual Reality Simulation Training. | 2016 | | | Denmark | | | | | Mean 3.2 months; range 2.4-5.0 months | Mean 3.2 months; range 2.4-5.0 months | Crosses 30-89 and >=90 day windows | SAME | YES | NO | Systematic review, nonpooled | ROBINS-I | Serious | Methods (PDF pp. 1-4): nonrandom distributed/massed cohorts; 36/42 returned; schedule assignment and baseline experience leave serious confounding. |
| STUDY_018 | Akhtar K; Sugand K; Wijendra A; Sarvesvaran M; Sperrin M; Standfield N; Cobb J; Gupte C | The Transferability of Generic Minimally Invasive Surgical Skills: Is There Crossover of Core Skills Between Laparoscopy and Arthroscopy? | 2016 | Journal of Surgical Education | 10.1016/j.jsurg.2015.10.018 | United Kingdom | prospective single-blinded crossover randomized controlled trial | 72 medical students randomized to 4 groups (2 control n=16 each; 2 training n=20 each) | VR LapMentor laparoscopic cholecystectomy or ArthroMentor knee arthroscopy training | matched no-training controls on same initial task | 1 week | 7 days | 7-29 days | TRANSFER | YES | NO | Systematic review, nonpooled | RoB 2 | Some concerns | Methods/Results (PDF pp. 1, 3-6): randomized four-group crossover; all 72 completed; multiple metrics with no prespecified delayed endpoint. |
| STUDY_019 | Ciechanski, Patrick | Effects of Transcranial Direct-Current | 2017 | | | Canada | | | | | 6 weeks (42 days) | 6 weeks (42 days) | 30-89 days | SAME | YES | NO | Systematic review, nonpool | RoB 2 | Some concerns | Methods/Results (PDF pp. 1, 3-6): concealed double-blind |

| | | | | | | | | | | | | | | | | | | | | |
|---|---|---|---|---|---|---|---|---|---|---|---|---|---|---|---|---|---|---|---|---|
| | | Stimulation on Neurosurgical Skill Acquisition: A Randomized Controlled Trial. | | | | | | | | | | | | | | | ed | | | sham-controlled allocation; all 22 completed; multiple outcome/subgroup analyses. |
| STUDY_020 | Nemani, Arun | Convergent validation and transfer of learning studies of a virtual reality-based pattern cutting simulator. | 2018 | Surgical Endoscopy | 10.1007/s00464-017-5802-8 | United States | Randomized controlled trial | 11 analyzed (VR n=6; no-training control n=5) | | | 14 days | 14 days | 7-29 days | SAME for primary; later ex-vivo transfer should be treated separately | YES for same-task retention | YES | Primary meta-analysis | RoB 2 | Some concerns | PMC5809233 Methods/Results: 18 students randomized 6/group; two-week retention; manual FLS scoring and several candidate outcomes. |
| STUDY_021 | Cychosz CC; Tofte JN; Johnson A; Gao Y; Phisitkul P | Fundamentals of Arthroscopic Surgery Training Program Improves Knee Arthroscopy Simulator Performance in Arthroscopic Trainees | 2018 | Arthroscopy | [DOI not printed in supplied PDF] | United States | prospective randomized trial | 43 medical students (FAST n=22; control n=21) | 5 self-guided FAST VR arthroscopy modules after baseline diagnostic knee arthroscopy | video/orientation plus no FAST training control | FAST 8.9 days vs control 9.5 days | Mean ~8.9 vs 9.5 days; assessment 1-2 weeks later | 7-29 days | TRANSFER | YES | YES | Primary meta-analysis | RoB 2 | Some concerns | Cychosz 2018 Methods/Tables 3-4 (PDF pp. 2-6) plus 2019 companion: randomized 22/21; no losses; automated metrics; nonblinded study. |
| STUDY_022 | LeBel Marie-Eve; Haverstock John; Cristancho Sayra; van Eimeren Lucia; Buckingham Gavin | Observational Learning During Simulation-Based Training in Arthroscopy: Is It Useful to Novices? | 2018 | Journal of Surgical Education | 10.1016/j.jsurg.2017.06.005 | Canada | Prospective randomized 3-group simulation study; 90 final-year medical students/very early PGY1 surgical residents, no prior arthroscopy/endoscopy/VR; p1-2. | | VR knee arthroscopy simulator; expert-video vs non-expert-video vs no-video control; same diagnostic arthroscopy task; randomized by coin toss; p1,3-4. | | Pretest, posttest, Test 3 at 1 week (plus Tests 2 and 4-5); p1,4. | 1 week | 7-29 days | SAME | YES | NO | Systematic review, nonpooled | RoB 2 | Some concerns | Methods/Results (PDF pp. 1-4): coin-toss allocation; approximately 28/28/26 available at retention; objective metrics and multiple comparisons. |
| STUDY_023 | Linsk Ali M; Monden Kimberley R; Sankaranarayanan Ganesh; Ahn Woojin; Jones Daniel B; De Suvranu; Schwaitzberg Steven D; Cao Caroline G L | Validation of the VBLaST pattern cutting task: a learning curve study | 2018 | Surgical Endoscopy | 10.1007/s00464-017-5895-0 | United States | Mixed experimental randomized 3-condition study; 24 medical students initially 10/group, final control n=9, FLS n=8, VBLaST n=7 after attrition; little/no prior surgery/simulator; p1-3. | 24 analyzed at delayed follow-up | FLS physical box trainer vs VBLaST-PC VR vs no-training control; 15 x 30-min sessions over 3 weeks; pattern-cutting task; p1-4. | | Pretest day 1, posttest day 15, retention day 16/2 weeks after last training session; Table 1 p4 and abstract p1. | 14 days | 7-29 days | SAME | YES | YES | Primary meta-analysis | RoB 2 | High | Methods/Results/Figures 2-7 (PDF pp. 2-10): 30 recruited, 24 analyzed; outliers were replaced by group means; small delayed cells. |
| STUDY_024 | Sugand Kapil; Wescott Robert A; Carrington | Training and Transfer Effect of FluoroSim, an | 2019 | Journal of Bone and Joint Surgery America | 10.2106/JBJS.18.00928 | United Kingdom | Single-blinded randomized controlled trial; 45 undergraduate medical students | | FluoroSim augmented-reality fluoroscopic DHS guidewire simulator: training | | TraumaVision baseline then final after 1-week washout and training | 7 days to first post-washout attempt | 7-29 days | SAME | YES only for first attempt after washout; | NO | Systematic review, nonpooled | RoB 2 | Some concerns | Methods/Tables I-III (PDF pp. 4, 6-8): electronic randomization and blinded |

| | | | | | | | | | | | | | | | | | | | | |
|---|---|---|---|---|---|---|---|---|---|---|---|---|---|---|---|---|---|---|---|---|
| | Richard; Hart Alister; van Duren Bernard H | Augmented Reality Fluoroscopic Simulator for Dynamic Hip Screw Guidewire Insertion: A Single-Blinded Randomized Controlled Trial | | n | | | unfamiliar with DHS/orthopaedic simulation; training n=23, control n=22; p1,4,7. | | 5 attempts week 1 + 1-week washout + 5 attempts week 2; control 1 attempt each week; both TraumaVision baseline/final transfer tests; p2,4,6. | | period; genuine post-washout first FluoroSim attempt = attempt 6; final formal comparison attempt 10 after extra practice; p4,6-8. | | | | reported formal between-group endpoint contaminated by additional practice | | | | | testing; complete primary follow-up; multiple endpoints/timepoints. |
| STUDY_025 | Sankaranarayanan Ganesh; Odlozil Coleman A; Wells Katerina O; Leeds Steven G; Chauhan Sanket; Fleshman James W; Jones Daniel B; De Suvranu | Training with cognitive load improves performance under similar conditions in a real surgical task | 2020 | The American Journal of Surgery | 10.1016/j.amjsurg.2020.02.002 | United States | Randomized 3-group study; 11 third-year medical students without prior FLS (control n=4, VR n=3, VR+cognitive load n=4); p1-2. | | Gen2-VR peg transfer; control no training vs VR training vs VR + two-digit multiplication cognitive load during last 100/150 trials; transfer = running 150-cm pig intestine under cognitive load; p1-2. | | Pretest, posttest after 15 sessions/3 weeks, retention 2 weeks later without interim practice; transfer immediately after retention; p1-2. | 14 days | 7-29 days | SAME | YES | NO | Systematic review, nonpooled | RoB 2 | Some concerns | Methods/Results (PDF pp. 1-8): randomization stated without concealment; very small groups; blinded transfer ratings but several figure-based outcomes. |
| STUDY_026 | Alvarez-Lopez, Fernando | Use of a Low-Cost Portable 3D Virtual Reality Simulator for Psychomotor Skill Training in Minimally Invasive Surgery: Task Metrics and Score Validity. | 2020 | Journal of Medical Internet Research | 10.2196/19723 | Spain | Prospective validation/retention cohort | 29 randomly selected for six-month retest | | | ~6 months | ~6 months | >=90 days | SAME | YES | NO | Systematic review, nonpooled | JBI cohort checklist (condensed items) | No global score | PubMed PMID 33107833: prospective validation cohort; 29 selected for six-month retest; selection and confounding not fully addressed. |
| STUDY_027 | Vamadevan, Anishan | Variable practice is superior to self-directed training for laparoscopic simulator training: a randomized trial. | 2024 | Surgical Endoscopy | 10.1007/s00464-024-10688-z | United Kingdom | Randomized trial | 36 randomized | | | 3-5 weeks | 3-5 weeks | Crosses 7-29 and 30-89 day windows | SAME / procedural | NO | NO | Systematic review, nonpooled | RoB 2 | Some concerns | PubMed PMID 38321334: randomized n=36 with 3-5-week retention; concealment, blinding, and missing-data handling incompletely reported. |
| STUDY_028 | Guseila Loredana M; Saranathan Archana; Jenison Eric L; Gil Karen M; Elias John J | Using virtual reality to maintain surgical skills during periods of robotic surgery | 2014 | Journal of Robotic Surgery | 10.1007/s11701-014-0465-0 | United States | Prospective repeated-measures maintenance cohort; 12 residents with no prior formal robotic training enrolled, 11 analyzed after | | Initial robot training to proficiency on needle passage, running suture pod, rocking peg board; then only biweekly virtual | | Week 0 proficiency and tissue-closure evaluation; week 8 robot evaluation after 8-week inactivity with | 8 weeks | 30-89 days | SAME / related robotic skill maintenance | NO | NO | Systematic review, nonpooled | JBI cohort checklist (condensed items) | No global score | Methods/Results (PDF pp. 1-6): 12 enrolled, 11 analyzed; all received scheduled maintenance practice; |

| | | | | | | | | | | | | | | | | | | | | |
|---|---|---|---|---|---|---|---|---|---|---|---|---|---|---|---|---|---|---|---|---|
| | | inactivity | | | | | one dropout; Ob/Gyn/urology/general surgery; Table 1 p2. | | robotic practice weeks 2,4,6; no clinical robot access; final robot evaluation week 8; no comparator; pp1-4. | | virtual practice weeks 2,4,6; Table 2 p2, methods pp3-4. | | | | | | | | | objective robot/simulator metrics. |
| STUDY_029 | Blumstein Gideon; Zukotynski Brian; Cevallos Nicolas; Ishmael Chad; Zoller Steven; Burke Zach; Clarkson Samuel; Park Howard; Bernthal Nicholas; SooHoo Nelson F | Randomized Trial of a Virtual Reality Tool to Teach Surgical Technique for Tibial Shaft Fracture Intramedullary Nailing | 2020 | Journal of Surgical Education | 10.1016/j.jsurg.2020.01.002 | United States | Blinded randomized prospective study; 20 first/second-year medical students with no prior tibial IMN/SawBones/VR gaming; VR n=10, standard guide n=10; 17 completed phase II; pp1-3. | | Osso VR tibial IMN tutorial/test vs printed surgical guide; SawBones tibial IMN evaluation immediately and 2 weeks later; no access/practice between phases; pp1-4. | | Phase I immediately post-training; phase II 2 weeks later; pp1,3-4. | 2 weeks | 7-29 days | SAME / transfer depending assessment platform | NO | NO | Systematic review, nonpooled | RoB 2 | Some concerns | Methods/Results (PDF pp. 2-7): sealed-envelope randomization and blinded evaluator; 17/20 reached delayed phase; booster exposure preceded later testing. |
| STUDY_030 | Lamblin Géry; Thiberville Gabriel; Druette Loic; Moret Stéphanie; Couraud Sébastien; Martin Xavier; Dubernard Gil; Chene Gautier | Virtual reality simulation to enhance laparoscopic salpingectomy skills | 2020 | Journal of Gynecology Obstetrics and Human Reproduction | 10.1016/j.jogoh.2020.101685 | France | Single-center prospective longitudinal study; 26 junior gynecology-obstetrics fellows (<4 semesters), all laparoscopic novices and no prior in-vivo salpingectomy; all completed; pp1-2. | | LapSim VR salpingectomy, 3 trials in session 1 and 3 in session 2; senior surgeon guidance on trial 2 each session; between sessions fellows performed in-vivo salpingectomy during hospital training; pp2,5-6. | | Session 1 Jan/Feb 2017 and session 2 May/Jun 2017, 3-month interval; pp1-2. | 3 months | >=90 days / approximately 90 days | SAME | YES | NO | Systematic review, nonpooled | JBI cohort checklist (condensed items) | No global score | Methods/Results (PDF pp. 1-6): all 26 completed; 84.6% had interim in-vivo exposure; single unblinded senior-surgeon rating. |
| STUDY_031 | Khan, Montaha W | Laparoscopic Skills Maintenance: A Randomized Trial of Virtual Reality and Box Trainer Simulators | 2014 | | | Australia | | | | | 1, 3, and 6 months | 1, 3, and 6 months | 30-89 and >=90 days | SAME | NO | NO | Systematic review, nonpooled | RoB 2 | High | Methods/Results (PDF pp. 1-5): sealed-envelope randomization; 26/41 completed all follow-ups and noncompletion related to lower performance. |
| STUDY_032 | Sinha Prashant; Hogle Nancy J; Fowler Dennis L | Do the laparoscopic skills of trainees deteriorate over time? | 2008 | Surgical Endoscopy | 10.1007/s00464-008-9929-5 | United States | Prospective single-cohort longitudinal study; 33 PGY1-3 general-surgery residents; all trained to criteria on LapSim and completed follow-up; pp1-3. | 33 residents | LapSim VR training to pass seven technical tasks (camera navigation, instrument navigation, coordination, grasping, lifting/grasping, cutting, clip applying); no required practice during 6 months; retest 3 attempts, first | | Baseline after proficiency and 6-month retest; retest subsequent two exams compared with baseline; pp1,3. | 6 months | >=90 days | SAME | NO / uncertain due simulator availability and acclimatization attempt | NO | Systematic review, nonpooled | JBI cohort checklist (condensed items) | No global score | Methods/Results (PDF pp. 1-6): all 33 completed six-month assessment; possible interim simulator access and acclimatization attempt. |

| | | | | | | | | | | | | | | | | | | | | |
|---|---|---|---|---|---|---|---|---|---|---|---|---|---|---|---|---|---|---|---|---|
| | | | | | | | | | acclimatization; pp1-3. | | | | | | | | | | | |
| STUDY_ 033 | Frithioff, Andreas | Automated summative feedback improves performanc e and retention in simulation training of mastoidecto my | 20 22 | Journal of Laryngol ogy & Otology | 10.1017/S002221512100 3352 | Denmar k | Randomized controlled trial | 24 randomized | | | 2-3 months | 2-3 months | Potentiall y crosses 30-89 and >=90 days dependin g exact follow-up dates | SAME | YES | NO | System atic review, nonpool ed | RoB 2 | Some concer ns | PubMed PMID 34709147: randomized n=24 with 2-3-month retention; concealment, assessor blinding, and complete delayed variance reporting unclear. |

#### Table S5. Primary endpoint decisions

| Study | Decision | Endpoint | Rationale |
|---|---|---|---|
| Chou 2006 | Included | OSATS quality score (maximum 35) | First eligible independent delayed objective contrast with recoverable mean, SD, and n. |
| Sabench Pereferrer 2013 | Included | Fine-dissection LapSim score (%) | First eligible independent delayed objective contrast with recoverable mean, SD, and n. |
| Jensen 2014 | Included | Lobectomy performance time with penalty | First eligible independent delayed objective contrast with recoverable mean, SD, and n. |
| Nemani 2018 | Included | VBLaST normalized pattern-cutting score | First eligible independent delayed objective contrast with recoverable mean, SD, and n. |
| Cychosz 2018 | Included | FAST composite arthroscopy score | First eligible independent delayed objective contrast with recoverable mean, SD, and n. |
| Linsk 2018 | Included | VBLaST normalized pattern-cutting score | First eligible independent delayed objective contrast with recoverable mean, SD, and n. |
| Zhang 2013 | Systematic review only | Not pooled | Delayed group means were graphical and error-bar type was not defined. |
| Van Bruwaene 2015 | Systematic review only | Not pooled | Delayed summaries were medians and interquartile ranges. |
| Akhtar 2016 | Systematic review only | Not pooled | Delayed multi-cohort summaries were median-based and lacked a compatible mean/SD contrast. |
| Sankaranarayanan 2020 | Systematic review only | Not pooled | Delayed means were graphical, dispersion was undefined, and cell sizes were very small. |

#### Statistical formulas

The pooled standard deviation was calculated as the square root of $\{[(nXR-1)SDXR^2 + (nC-1)SDC^2]/(nXR+nC-2)\}$. Cohen's d was the oriented mean difference divided by the pooled standard deviation. Hedges' g equaled Jd, where $J = 1 - 3/[4(nXR+nC)-9]$. The sampling variance equaled $J^2[(nXR+nC)/(nXR\ nC) + d^2/\{2(nXR+nC-2)\}]$.

Paule-Mandel $\tau^2$ was obtained as the nonnegative value for which the random-effects Q equaled $k-1$. The Hartung-Knapp confidence interval used a t distribution with $k-1$ degrees of freedom. Model-specific prediction intervals were calculated when at least three studies contributed. For the DL sensitivity, a conventional normal-theory diagnostic interval was recalculated for numerical QA but was not reported as a substantive result.

#### Table S6. Complete synthesis and sensitivity results

| Analysis | k | Method | Hedges' g | 95% CI | τ² | I², % | Prediction interval | Interpretation |
|---|---|---|---|---|---|---|---|---|
| Primary random-effects synthesis | 6 | Paule–Mandel random effects with Hartung–Knapp CI | 1.39 | −0.94 to 3.72 | 4.53 | 88.0 | −5.04 to 7.81 | Average effect is imprecise; between-study heterogeneity is substantial |
| Task relation: same | 3 | Paule–Mandel random effects with Hartung–Knapp CI | 2.95 | −2.94 to 8.84 | 4.96 | 86.8 | −30.26 to 36.16 | Exploratory sparse subgroup; prediction interval reported only in supplement |
| Task relation: transfer | 3 | Paule–Mandel random effects with Hartung–Knapp CI | 0.01 | −1.79 to 1.81 | 0.37 | 75.3 | −9.41 to 9.42 | Exploratory sparse subgroup; prediction interval reported only in supplement |
| Comparator: inactive/no training | 4 | Paule–Mandel random effects with Hartung–Knapp CI | 2.30 | −1.27 to 5.88 | 4.53 | 85.5 | −8.05 to 12.66 | Exploratory sparse subgroup; prediction interval reported only in supplement |
| Comparator: active trainer | 2 | Paule–Mandel random effects with Hartung–Knapp CI | −0.37 | −6.10 to 5.36 | 0.23 | 54.9 | Not calculated | Exploratory sparse subgroup; prediction interval reported only in supplement |
| Retention: 7–29 days | 3 | Paule–Mandel random effects with Hartung–Knapp CI | 2.77 | −3.76 to 9.31 | 6.24 | 90.3 | −34.36 to 39.91 | Exploratory sparse subgroup; prediction interval reported only in supplement |
| Retention: 30–89 days | 3 | Paule–Mandel random effects with Hartung–Knapp CI | 0.20 | −2.28 to 2.68 | 0.85 | 87.7 | −13.61 to 14.01 | Exploratory sparse subgroup; prediction interval reported only in supplement |
| DerSimonian–Laird, normal CI (same six studies) | 6 | DerSimonian–Laird random effects with normal CI | 1.16 | 0.11 to 2.22 | 1.39 | 88.0 | Not calculated | Conventional DL/normal sensitivity; prediction interval not reported in main table |
| Randomized trials only | 5 | Paule–Mandel random effects with Hartung–Knapp CI | 1.47 | −1.66 to 4.60 | 5.89 | 89.2 | −7.05 to 9.99 | Sensitivity analysis |
| Exclude figure-derived estimate | 5 | Paule–Mandel random effects with Hartung–Knapp CI | 0.61 | −0.76 to 1.99 | 1.02 | 82.2 | −2.97 to 4.20 | Sensitivity analysis |
| Exclude high/serious risk of bias | 4 | Paule–Mandel random effects with Hartung–Knapp CI | 0.47 | −1.52 to 2.46 | 1.32 | 81.2 | −5.15 to 6.09 | Sensitivity analysis |
| Leave-one-out: omit Chou 2006 | 5 | Paule–Mandel random effects with Hartung–Knapp CI | 1.68 | −1.31 to 4.66 | 5.35 | 90.1 | −6.44 to 9.79 | Leave-one-out sensitivity |
| Leave-one-out: omit Sabench Pereferrer 2013 | 5 | Paule–Mandel random effects with Hartung–Knapp CI | 1.47 | −1.66 to 4.60 | 5.89 | 89.2 | −7.05 to 9.99 | Leave-one-out sensitivity |
| Leave-one-out: omit Jensen 2014 | 5 | Paule–Mandel random effects with Hartung–Knapp CI | 1.84 | −0.85 to 4.52 | 4.23 | 83.7 | −5.40 to 9.07 | Leave-one-out sensitivity |
| Leave-one-out: omit Nemani 2018 | 5 | Paule–Mandel random effects with Hartung–Knapp CI | 1.24 | −1.80 to 4.27 | 5.63 | 89.0 | −7.08 to 9.55 | Leave-one-out sensitivity |
| Leave-one-out: omit Cychosz 2018 | 5 | Paule–Mandel random effects with Hartung–Knapp CI | 1.59 | −1.49 to 4.67 | 5.68 | 90.4 | −6.78 to 9.96 | Leave-one-out sensitivity |
| Leave-one-out: omit Linsk 2018 | 5 | Paule–Mandel random effects with Hartung–Knapp CI | 0.61 | −0.76 to 1.99 | 1.02 | 82.2 | −2.97 to 4.20 | Leave-one-out sensitivity |

#### Table S7. Study-level risk-of-bias judgments

| Study | Tool | D1/I1 | D2/I2 | D3/I3 | D4/I4 | D5/I5 | D6 | D7 | Overall |
|---|---|---|---|---|---|---|---|---|---|
| Chou DS 2006 | RoB 2 | Some concerns | Low | Low | Some concerns | Some concerns | | | Some concerns |
| Verdaasdonk EGG 2007 | RoB 2 | Some concerns | Low | Low | Low | Some concerns | | | Some concerns |
| Snyder CW 2010 | ROBINS-I | Moderate | Serious | Low | Moderate | Moderate | Low | Moderate | Serious |
| Kruglikova I 2010 | RoB 2 | Some concerns | Some concerns | Some concerns | Low | Some concerns | | | Some concerns |
| Mulla M 2012 | RoB 2 | Some concerns | Low | Some concerns | Low | Some concerns | | | Some concerns |
| Gallagher AG 2012 | RoB 2 | Some concerns | Low | Low | Low | Some concerns | | | Some concerns |
| Zhang L 2013 | RoB 2 | Some concerns | Some concerns | Some concerns | Some concerns | Some concerns | | | Some concerns |
| Sabench Pereferrer F 2013 | ROBINS-I | Serious | Moderate | Low | Moderate | Moderate | Low | Moderate | Serious |
| Jensen K 2014 | RoB 2 | Some concerns | Low | Some concerns | Low | Some concerns | | | Some concerns |
| Bjerrum F 2015 | RoB 2 | Some concerns | Low | High | Low | Some concerns | | | High |
| Van Bruwaene S 2015 | RoB 2 | Some concerns | Low | Low | Some concerns | Some concerns | | | Some concerns |
| Andersen SAW 2016 | ROBINS-I | Serious | Moderate | Low | Moderate | Moderate | Low | Moderate | Serious |
| Akhtar K 2016 | RoB 2 | Some concerns | Some concerns | Low | Low | Some concerns | | | Some concerns |
| Ciechanski P 2017 | RoB 2 | Low | Low | Low | Low | Some concerns | | | Some concerns |
| Nemani A 2018 | RoB 2 | Some concerns | Low | Some concerns | Some concerns | Some concerns | | | Some concerns |
| Cychosz 2018/2019 study family | RoB 2 | Some concerns | Low | Low | Low | Some concerns | | | Some concerns |
| LeBel ME 2018 | RoB 2 | Some concerns | Some concerns | Some concerns | Low | Some concerns | | | Some concerns |
| Linsk AM 2018 | RoB 2 | Some concerns | Some concerns | High | Low | Some concerns | | | High |
| Sugand K 2019 | RoB 2 | Low | Some concerns | Low | Low | Some concerns | | | Some concerns |
| Sankaranarayanan G 2020 | RoB 2 | Some concerns | Some concerns | Some concerns | Some concerns | Some concerns | | | Some concerns |
| Vamadevan A 2024 | RoB 2 | Some concerns | Some concerns | Some concerns | Some concerns | Some concerns | | | Some concerns |
| Blumstein G 2020 | RoB 2 | Low | Some concerns | Some concerns | Some concerns | Some concerns | | | Some concerns |
| Khan MW 2014 | RoB 2 | Low | Some concerns | High | Some concerns | Some concerns | | | High |
| Frithioff A 2022 | RoB 2 | Some concerns | Some concerns | Some concerns | Some concerns | Some concerns | | | Some concerns |

RoB 2 and ROBINS-I domain names are tool-specific. The Cychosz companion reports are represented by one risk-of-bias unit. Complete design-specific JBI appraisals are provided in Tables S10a-b.

#### Table S8. Revised study-selection accounting

| Stage | Count | Treatment in final PRISMA |
|---|---|---|
| Records identified | 8,628 | Identification |
| Duplicates removed | 3,573 | Before screening |
| Records screened | 5,055 | All deduplicated records underwent title/abstract screening |

| | | |
|---|---|---|
| Records excluded at record level | 4,122 | Manual title/abstract screening |
| Reports sought for retrieval | 933 | Full-text retrieval |
| Reports not retrieved | 0 | All sought reports were obtained |
| Reports assessed for eligibility | 933 | Full-text assessment |
| Full-text reports excluded | 899 | Did not meet prespecified population, intervention, outcome, design/report-type, or follow-up criteria |
| Reports included | 34 | 33 distinct studies |
| Distinct studies included | 33 | One companion-report pair linked as one study family |
| Primary quantitative synthesis | 6 | Independent study-family contrasts |

#### Table S9. GRADE certainty of evidence

| Outcome | Studies | Participants | Effect | Certainty | Reasons |
|---|---|---|---|---|---|
| First eligible uncontaminated delayed objective technical performance after XR versus comparator | 6 | 162 selected-arm participants | Hedges' g 1.39 (95% CI -0.94 to 3.72); PI -5.04 to 7.81 | Very low | Very serious risk of bias; very serious inconsistency; serious indirectness; very serious imprecision. Publication bias not formally assessed (k=6). |

#### Table S10a. JBI cohort item-level appraisal

| Study | Inclusion criteria | Exposure reliable | Confounding addressed | Outcome reliable | Follow-up handled | Rationale |
|---|---|---|---|---|---|---|
| Ogan K 2004 | Yes | Yes | No | Yes | Yes | Prospective longitudinal cohort with an expertise-reference group; the reference group is not a causal control. |
| Stefanidis D 2005 | Yes | Yes | No | Yes | Yes | Single-arm repeated-measures cohort with complete delayed follow-up. |
| Maagaard M 2011 | Yes | Yes | No | Yes | Unclear | Six- and 18-month follow-up cohort with expert reference; one novice was lost at 18 months. |
| Alvarez-Lopez F 2020 | Unclear | Yes | Unclear | Yes | Unclear | Prospective validation/retention cohort; full report was unavailable in the supplied archive. |
| Sinha P 2008 | Yes | Yes | Unclear | Yes | Unclear | Prospective single-cohort retention study; interim access and acclimatization may contaminate follow-up. |

#### Table S10b. JBI quasi-experimental item-level appraisal

| Study | Q1 | Q2 | Q3 | Q4 | Q5 | Q6 | Q7 | Q8 | Q9 | Rationale |
|---|---|---|---|---|---|---|---|---|---|---|
| Windsor JA | Yes | N/A | N/A | No | No | Yes | Yes | Yes | Yes | Repeated acquisition, loss, and reacquisition measurements in one trained group. |

| | | | | | | | | | | |
|---|---|---|---|---|---|---|---|---|---|---|
| 2005 | | | | | | | | | | |
| Janse JA 2013 | Yes | N/A | N/A | No | No | Unclear | Yes | Yes | Yes | Prospective repeated-measures training study; experts are a validation reference, not a causal control. |
| Guseila LM 2014 | Yes | N/A | N/A | No | No | Unclear | Yes | Yes | Unclear | One-group maintenance intervention with scheduled biweekly practice and no concurrent control. |
| Lamblin G 2020 | Yes | N/A | N/A | No | No | Yes | Yes | Unclear | Yes | One-group before-after study with interim clinical exposure and incompletely documented rating reliability. |

JBI quasi-experimental questions: Q1 clear cause/effect; Q2 participants in comparisons similar; Q3 similar care apart from the exposure; Q4 control group; Q5 multiple measurements before and after; Q6 follow-up complete/handled; Q7 outcomes measured similarly; Q8 outcomes measured reliably; Q9 appropriate statistical analysis. N/A was used for between-group comparability items in single-group repeated-measures designs. No global score was assigned for either tool.

#### Table S11. PRISMA 2020 checklist updates for this pass

| Item | Requirement | Location/status |
|---|---|---|
| 6 | Information sources and last search date | Methods and Table S1; cutoff May 5, 2026 |
| 7 | Full search strategies | Table S1 and accompanying workbook; reconstructed yields explicitly labeled |
| 8 | Selection process | Methods: all 5,055 deduplicated records treated as screened; two-reviewer process stated |
| 11 | Risk of bias | Methods; Figure 3; complete JBI table in this supplement |
| 13 | Synthesis methods | Methods: estimand, endpoint hierarchy, multi-arm and independence rules |
| 15 | Certainty assessment | Methods and Table S9; GRADE |
| 16 | Study selection | Results and Figure 1; 4,122 record-level exclusions |
| 22 | Certainty | Results, Discussion, and Table S9: very low |
